# Neonatal meconium reveals concurrent microplastic and metal exposure in an urban South Asian birth cohort

**DOI:** 10.64898/2026.05.12.26352974

**Authors:** Ifthikhar Zaman, Mahdi Muhammad Moosa, Eshra Sultana, Rownok Ara Sara, Nushrat Jahan, Sukanna Mysha, Nazifa Tabassum Tasnim, Mohammad Moniruzzaman, Md Yasir Arafat, M. Mahboob Hossain, Nadia Sultana Deen

## Abstract

Neonatal meconium provides a non-invasive matrix for assessing prenatal/near-birth exposure to environmental contaminants. Although microplastics and metals have each been reported in human biological samples, integrated assessments of concurrent particle and metal exposure in meconium remain scarce, particularly in South Asia. In this cross-sectional biomonitoring study, meconium from 30 Cesarean-delivered neonates born in Dhaka, Bangladesh, was analyzed for microplastic occurrence, morphology, and polymer composition using stereomicroscopy, scanning electron microscopy, and Raman spectroscopy, and for fifteen metals using inductively coupled plasma mass spectrometry along with exploratory meconium bacterial profiles. Maternal breast milk from a subset of lactating mothers was analyzed as a complementary maternal exposure context. Microplastics were detected in all analyzable meconium samples (n=28), with a median burden of 149 particles/g wet weight, dominated by polyethylene terephthalate fragments and nylon fibers; most fragments were below 200 µm. Breast milk likewise contained microplastics in all analyzed samples, indicating a potential postnatal exposure route. All fifteen measured metals were detected in every analyzable meconium sample, with median Pb and Cr concentrations of 1.18 and 3.92 µg/g dry weight, respectively. Selenium and copper concentrations declined significantly with gestational age. No microplastic-element associations remained significant after multiple-testing correction, providing no evidence of a consistent relationship between total particle burden and elemental concentrations in this cohort. Integrating 16S profiling that revealed low-complexity, heterogeneous bacterial communities, this study provides one of the first combined meconium-based assessments of microplastic, metal, and microbial signals from the region and highlights meconium as a practical matrix for early-life biomonitoring.

## 1. Introduction

Microplastics (MPs), defined as plastic particles <5 mm, are ubiquitous environmental contaminants increasingly detected in human biological samples, including feces, blood, placenta, and breast milk, indicating that human exposure is widespread and that plastic particles or polymer-derived signals may enter internal biological compartments (Leslie et al., 2022; Ragusa et al., 2022, 2021; Schwabl et al., 2019). Neonates are particularly vulnerable to environmental contaminants, plausibly because early development is characterized by rapid tissue growth, immature barrier and immune functions, and impaired detoxification and clearance pathways (Landrigan and Goldman, 2011).

Meconium, the first neonatal stool, begins forming in the fetal gastrointestinal tract at approximately 11-14 weeks of gestation and accumulates fetal gastrointestinal contents over an extended prenatal window (Mitchell and Chandraharan, 2018). This makes it a useful non-invasive matrix for assessing prenatal exposure to environmental contaminants (Michelsen-Correa et al., 2021). While cord blood and urine generally reflect more recent or circulating exposure, meconium can provide a more integrated record of fetal exposure across a longer prenatal window. Recent studies from the USA and China have reported MPs or plastic polymers in meconium, placenta, cord blood, infant feces, and breast milk, indicating that fetal and early neonatal exposure to plastic particles or polymer-derived contaminants is biologically plausible (Liu et al., 2023; Zhang et al., 2021; Zhu et al., 2024). However, comparable neonatal biomonitoring data from South Asia remain limited.

Meconium is also an established matrix for prenatal heavy metal biomonitoring (Michelsen-Correa et al., 2021). This is particularly relevant in densely urbanized environments where plastic pollution, traffic emissions, industrial activities, contaminated waterways, and legacy waste sites may contribute to complex early-life chemical exposures. MPs can adsorb trace metals and other contaminants under environmental conditions, raising concern that they may contribute to combined particle-chemical exposure through a proposed “Trojan horse” mechanism (Carriera et al., 2023; Holmes et al., 2012). However, the biological relevance of this mechanism in human prenatal exposure remains uncertain, and studies pairing MP characterization with metal profiling in the same neonatal exposure matrix remain scarce.

Dhaka, the capital of Bangladesh, provides an important setting for addressing this gap because it combines high population density, extensive plastic use, poor plastic waste management, traffic-related pollution, industrial contamination of waterways, and legacy tannery-associated metal pollution (Hossain et al., 2021; Parvin et al., 2022; Tanvir et al., 2022). This combination of particle and chemical exposures makes Dhaka a relevant urban context in which to assess whether neonates are exposed to MPs and metals concurrently. In this study, we characterized MPs, including their occurrence, morphology, and polymer composition in meconium from a neonatal cohort in Dhaka. Additionally, we quantified fifteen metals in overlapping meconium samples to assess concurrent prenatal or near-birth co-exposure. Maternal breast milk samples were also analyzed from a subset of lactating mothers who consented, providing complementary maternal context. We further explored associations between meconium MP burden, metal concentrations, neonatal demographic factors, and pilot meconium bacterial profiles generated from a subset of samples. To our knowledge, this represents one of the first integrated assessments of MPs and metals in neonatal meconium from an urban South Asian birth cohort.

## 2. Methods

### 2.1. Study Design and Ethics

This cross-sectional biomonitoring study was conducted at Ad-Din Medical College Hospital, Dhaka, Bangladesh (August-October 2025). Thirty Cesarean-delivered neonates (gestational age ≥34 weeks, no major congenital malformations) were enrolled. The study was approved by the BRAC University Institutional Review Board (Approval no: BRACUIRB120240014) and conducted per the Declaration of Helsinki. Written informed consent was obtained from mothers before sample collection.

### 2.2. Sample Collection

First-pass meconium (n = 30) was collected within 0-24 h of birth into pre-cleaned sterile glass vials and stored at -20 °C. Breast milk samples (n = 20) were collected from consenting lactating mothers by manual expression. All containers were pre-cleaned and verified free of microplastic contamination; non-plastic materials (glass and metal) were used throughout. Two meconium samples were lost during microplastic processing, and two yielded insufficient volume for elemental analysis, resulting in 28 samples available for individual analyses and 26 for combined MP-metal analyses. A detailed method is provided in Supplementary Materials 1.

### 2.3. Questionnaire Administration

A structured questionnaire was administered to parents or legal guardians at the time of sample collection. It comprised five sections: (A) neonatal information (sex, date of birth, gestational age), (B) parental demographics, (C) infant feeding practices (exclusive breastfeeding, formula feeding, or mixed feeding, including formula brand), (D) neonatal health status (including clinically diagnosed or suspected septicemia), and (E) use of infant care products (soaps, lotions, wipes, diapers, with brand identification where available). These variables were recorded as cohort/contextual information for analysis as appropriate.

### 2.4. Microplastic Isolation and Identification

Microplastic extraction was performed on meconium (0.5 g) and breast milk (1 mL). Organic matter was digested using Fenton oxidation (30% H₂O₂ with Fe²⁺ catalyst, 60 °C, 48 h). Density separation used saturated NaCl (∼1.2 g/cm³), followed by vacuum filtration through glass microfiber filters (0.10 µm). Particles were enumerated under stereomicroscopy (40×) and classified by morphology (fragment or fiber) and size. Procedural blanks were processed alongside each batch; counts were blank-corrected. Polymer identification used macro-Raman spectroscopy (HORIBA MacroRAM, 400–1800 cm⁻¹) matched via the OpenSpecy platform (Cowger et al., 2021), with assignments accepted at an HQI ≥45%. Selected particles were examined by scanning electron microscopy (JEOL JSM-7610F). Contamination control procedures, density separation limitations, and additional methodological details are provided in Supplementary Materials 1.

### 2.5. Elemental Analysis

Fifteen trace elements (As, Se, Pb, Cd, Cr, Ni, Cu, Zn, Hg, Mn, Co, Al, Be, V, and Fe) were quantified in meconium by inductively coupled plasma mass spectrometry (ICP-MS; PerkinElmer NexION 2000). Dried, homogenized meconium (0.1-0.5 g) was digested using microwave-assisted acid digestion adapted from US EPA Method 3051A, with 65% ultra-pure nitric acid and 30% hydrogen peroxide (7:3, v/v) at 200 °C for 30 min (Milestone Start D, Italy). Digested solutions were diluted to 25 mL with deionized water, filtered, and analyzed. Calibration used ICP-multi-element standard solution XIII (Merck, Germany), and internal standards (⁴⁵Sc, ⁷³Ge, ⁸⁹Y, ¹¹⁵In, ¹⁸⁵Re, ¹⁹³Ir, ²⁰⁹Bi) were added to correct for instrumental drift and matrix effects. Accuracy was verified using certified reference materials (SRM NIST 1648a and SRM 955d), with recoveries ranging from 91.8% to 108.3%. Reagent blanks, filter blanks, and spike recoveries were performed as part of quality control. Full analytical details, limits of detection, and precision data are provided in Supplementary Materials 1.

### 2.6. Meconium Microbial Community Analysis

Twelve meconium samples were included in the exploratory microbial analysis. The combined subset contained six male and six female neonates, with three high- and three low-MP samples within each sex. Given the small sample size, this analysis was considered hypothesis-generating. Genomic DNA was extracted using the DNeasy PowerSoil Pro Kit (QIAGEN). Quality and concentration were assessed by spectrophotometry and agarose gel electrophoresis. Full-length 16S rRNA gene amplicons (∼1,465 bp) were generated using primers 27F and 1492R, spanning the V1-V9 regions. Libraries were prepared with the Native Barcoding Kit (SQK-NBD114.96) and sequenced on the Oxford Nanopore MinION Mk1C.

Raw Oxford Nanopore 16S amplicon FASTQ files were inspected using SeqKit for read counts and length distributions (Shen et al., 2016). Reads were filtered to retain near-full-length 16S amplicons using a 1400-1700 bp length window. Filtered reads were taxonomically profiled using Emu (Curry et al., 2022) with the Emu 2026 database (Dutton, 2026).

For visualization and downstream summaries, Emu abundance outputs were collapsed to the genus level by summing relative abundances of all species assigned to the same genus. For the stacked relative-abundance plot, the eight most abundant genera across the combined dataset were shown individually, and remaining genera were grouped as “Other.” Shannon diversity was calculated from the complete genus-level relative-abundance profile of each sample rather than from the reduced plotting table. Genus-level Bray-Curtis dissimilarity, principal coordinates analysis, and exploratory associations with available clinical and exposure variables were evaluated as described in Supplementary Methods S1.5.

### 2.7. Statistical Analysis

Except for the microbiome analyses described in Section 2.6 and Supplementary Methods S1.5, all analyses were performed in Python (v3.14; SciPy v1.17.1, pandas v3.0.2). Normality was assessed using the Shapiro–Wilk test, and non-parametric methods were used throughout. Meconium MP counts were normalized to 1 g wet weight; breast milk counts are reported per mL. Samples exceeding the counting ceiling (500 fragments per 0.5 g) were conservatively recorded at that value. Spearman correlations assessed MP-metal associations (n = 26) and gestational age relationships. Mann-Whitney U tests compared groups by sex and MP burden. Benjamini–Hochberg false-discovery-rate correction was applied to analyses involving 15 parallel metal comparisons, including MP-metal, sex-metal, and gestational-age-metal analyses. Paired mother-neonate MP comparisons used Spearman correlations on 19 valid pairs. A two-tailed p < 0.05 was considered significant. Full statistical analysis details are provided in Supplementary Materials 1.

## 3. Results and Discussion

### 3.1 Demographic and clinical characteristics

The study cohort comprised 30 neonates delivered by Cesarean section at Ad-Din Medical College Hospital, Dhaka, between August and October 2025. Nineteen neonates (63.3%) were male and eleven (36.7%) female, with a median gestational age of 37.0 weeks (IQR: 36.1-38.2; range: 34.3-40.0). Nineteen (63.3%) were born at term (≥37 weeks), while eleven (36.7%) were classified as late preterm (34-36 weeks). Regarding feeding practices at the time of enrollment, fifteen neonates (50.0%) were exclusively breastfed, thirteen (43.3%) received mixed feeding combining breast milk and formula, and two (6.7%) were exclusively formula-fed. Infant care products were divided between traditional cotton wraps known locally as katha (n = 17, 56.7%) and disposable diapers (n = 13, 43.3%). All participants resided within the Dhaka metropolitan area (Fig. S2), a megacity of over 22 million inhabitants characterized by extensive plastic waste mismanagement, industrial contamination of surrounding waterways, and heavy reliance on packaged food and drinking water (Hossain et al., 2021, 2020; Parvin et al., 2022).

The high proportion of mixed feeding (43.3%) aligns with national survey data indicating that approximately 55% of Bangladeshi infants under six months are exclusively breastfed (BUHN, n.d.). This feeding pattern is relevant to microplastic exposure assessment because mixed-fed neonates receive microplastics through both breast milk and formula routes, and formula preparation using polypropylene feeding bottles has been shown to release millions of microplastic particles per feed (Li et al., 2020; Liu et al., 2023). The distinction between katha and disposable diapers is also noteworthy as a potential differential source of postnatal microplastic exposure, given that disposable diapers contain synthetic polymers, including polypropylene and polyethylene in their outer layers, absorbent core, and adhesive components, whereas katha is a traditional cotton-based cloth unlikely to contribute synthetic microplastic fibers. However, because meconium accumulates from approximately the twelfth week of gestation and primarily reflects prenatal exposure, neither feeding mode nor infant care product type was analyzed as an explanatory variable for meconium contaminant burden in this study.

The inclusion of eleven late preterm neonates (34-36 weeks) alongside nineteen term neonates enables exploratory comparison across gestational ages, although the unequal group sizes and narrow gestational age range (34.3-40.0 weeks) limit the statistical power of such comparisons. All deliveries were by Cesarean section, which is relevant for the microbiome analysis as it excludes direct vaginal microbial inoculation during birth.

### 3.2 Neonatal meconium microplastics

Microplastics were detected in all 28 analyzable meconium samples (100% detection; two samples were lost during processing). After normalization to 1.0 g wet weight, fragments were the dominant particle type, with a median count of 134 particles/g (IQR: 51.5-276.5; range: 26 to ≥1000). Two samples exceeded the counting ceiling of 500 fragments per 0.5 g and were conservatively recorded at that value, such that the highest counts represent minimum estimates. Fibers were detected in 25/28 samples (89.3%), with a median of 8 fibers/g (IQR: 4-11). The median total microplastic burden was 149 particles/g (IQR: 58-294.5) (Fig. 1A; Table 1).

**Fig. 1:**
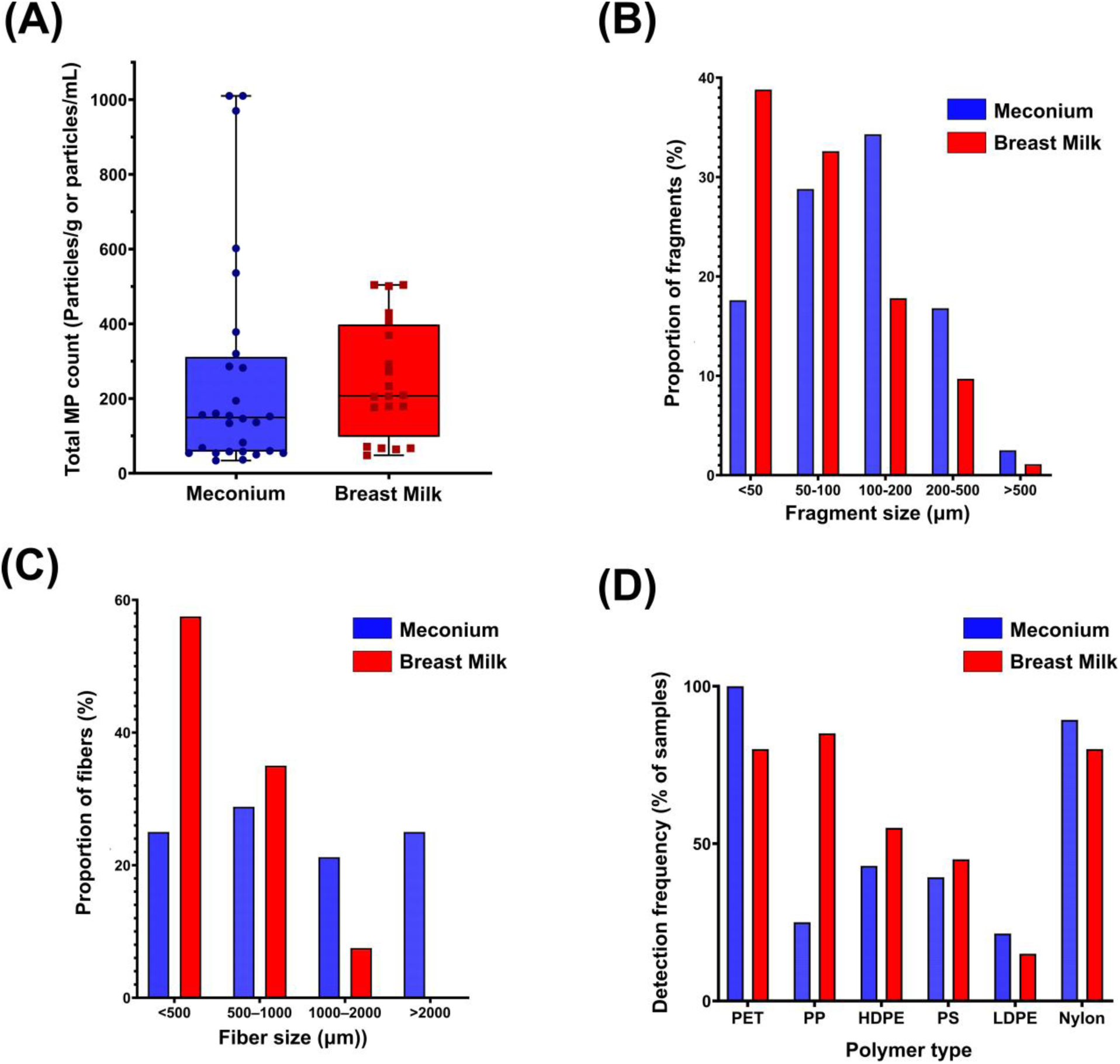
Microplastic burden and characteristics in neonatal meconium and maternal breast milk. **(A)** Total microplastic counts are shown as box-and-whisker plots with individual data points. Meconium values are expressed as particles/g wet weight, and breast milk values as particles/mL; matrices are shown side-by-side for descriptive context only because units differ. Two meconium samples exceeded the fragment-counting ceiling; their total burdens were recorded as ≥1010 particles/g. **(B)** Fragment size distribution by matrix (meconium: *n* = 358; breast milk: *n* = 258). **(C)** Fiber length distribution by matrix (meconium: *n* = 104; breast milk: *n* = 40). **(D)** Detection frequency of polymer types identified by Raman spectroscopy. PET, polyethylene terephthalate; PP, polypropylene; HDPE, high-density polyethylene; PS, polystyrene; LDPE, low-density polyethylene; nylon, polyamide.

**Table 1.** Summary of microplastic burden and polymer detection in neonatal meconium and maternal breast milk. Microplastic count data are presented as medians with interquartile ranges (IQR) and ranges. Meconium values are expressed as counts/g wet weight, and breast milk values as counts/mL. Polymer rows indicate the percentage of samples in which each polymer was detected. Male and female subgrouping refers to neonate sex; for breast milk, male and female indicate mothers of male and female neonates, respectively. p-values were calculated using Mann–Whitney U tests. PET, polyethylene terephthalate; PP, polypropylene; HDPE, high-density polyethylene; PS, polystyrene; LDPE, low-density polyethylene; nylon, polyamide.

| **Measure** | **n** | **Median** | **IQR** | **Range** | **p-value**<br>**(Male vs**<br>**Female)** |
| --- | --- | --- | --- | --- | --- |
| Number of samples | 28 |  |  |  |  |
| Male | 17 |  |  |  |  |
| Female | 11 |  |  |  |  |
| Detection rate | 28/28<br>(100%) |  |  |  |  |
| Total MP (count/g) |  |  |  |  |  |
| Overall | 28 | 149 | 58.0-294.5 | 34-<br>≥1010 |  |
| Male | 17 | 194 | 68.0-536.0 |  | 0.0454 |
| Female | 11 | 134 | 54.0-153.0 |  |  |
| Fragments (count/g) |  |  |  |  |  |
| Overall | 28 | 134 | 51.5-276.5 | 26-1000 |  |
| Male | 17 | 190 | 66.0-368.0 |  | 0.0629 |
| Female | 11 | 130 | 44.0-142.0 |  |  |
| Fibers (count/g) |  |  |  |  |  |
| Overall | 28 | 8 | 4.0-11.0 | 0-360 |  |
| Male | 17 | 6 | 4.0-10.0 |  | 0.6688 |
| Female | 11 | 8 | 7.0-12.0 |  |  |
| **Polymer detection (% of samples)** |  |  |  |  |  |
| PET | 100.0% |  |  |  |  |
| HDPE | 42.9% |  |  |  |  |
| PS | 39.3% |  |  |  |  |
| PP | 25.0% |  |  |  |  |
| LDPE | 21.4% |  |  |  |  |
| Nylon (fiber) | 89.3% |  |  |  |  |
| Number of samples | 20 |  |  |  |  |
| Mothers of male neonates | 12 |  |  |  |  |
| Mothers of female neonates | 8 |  |  |  |  |
| Detection rate | 20/20<br>(100%) |  |  |  |  |
| Total MP (count/mL) |  |  |  |  |  |
| Overall | 20 | 207 | 149.8-378.8 | 48-504 |  |
| Male | 12 | 206 | 67.0-413.2 |  | 0.8469 |
| Female | 8 | 220 | 178.2-277.0 |  |  |
| Fragments (count/mL) |  |  |  |  |  |
| Overall | 20 | 204 | 145.8-376.5 | 44-500 |  |
| Male | 12 | 204 | 66.8-409.5 |  | 0.9692 |
| Female | 8 | 216 | 172.8-276.0 |  |  |
| Fibers (count/mL) |  |  |  |  |  |
| Overall | 20 | 3 | 1.0-4.0 | 0-6 |  |
| Male | 12 | 2 | 0.8-4.0 |  | 0.2528 |
| Female | 8 | 4 | 3.0-4.0 |  |  |
| **Polymer detection (% of samples)** |  |  |  |  |  |
| PP | 85.0% |  |  |  |  |
| PET | 80.0% |  |  |  |  |
| HDPE | 55.0% |  |  |  |  |
| PS | 45.0% |  |  |  |  |
| LDPE | 15.0% |  |  |  |  |
| Nylon (fiber) | 80.0% |  |  |  |  |

Fragment sizes (n = 358) ranged predominantly within the small-particle fraction, with a mean of 143.2 ± 152.4 µm and a median of 100 µm; 81.0% of fragments were <200 µm (Fig. 1B, C; Fig. S1, S3). Fibers (n = 104) were substantially longer, with a mean length of 1301.8 ± 1057.8 µm and a median of 920 µm. These data indicate that meconium-associated MPs in this cohort were numerically dominated by small fragments, with fewer but longer nylon fibers.

Raman spectroscopy identified polyethylene terephthalate as the dominant fragment polymer, detected in all meconium samples (100%). Other fragment polymers included high-density polyethylene (HDPE; 42.9%), polystyrene (PS; 39.3%), polypropylene (PP; 25.0%), and low-density polyethylene (LDPE; 21.4%). All fibers were identified as nylon (polyamide) (Fig. 1D).

The 100–200 micrometer range accounted for 34.3% of all measured fragments, followed by 50-100 micrometers (28.8%), less than 50 micrometers (17.6%), 200-500 micrometers (16.8%), and greater than 500 micrometers (2.5%). This distribution is consistent with prior studies in which more than 80% of detected particles in meconium were in the 20–400 micrometer range (Liu et al., 2023; Zhu et al., 2024). This size profile is also compatible with proposed transplacental passage of smaller particles, although the present study did not determine the route by which particles entered meconium.

Fiber lengths were broadly distributed across all size categories: less than 500 micrometers (25.0%), 500-1000 micrometers (28.8%), 1000–2000 micrometers (21.2%), and greater than 2000 micrometers (25.0%). The presence of fibers exceeding 2000 micrometers in meconium is notable because particles of this length would be difficult to transport through placental tissue. The route by which such long fibers entered the meconium cannot be resolved from the present data. Sample S17 was a clear outlier with 360 fibers per gram, far exceeding the cohort median of 8 per gram. Excluding this sample, the mean fiber count decreased from 21.3 to 8.7 fibers per gram while the median remained unchanged at 8. Three samples (S24, S29, S30) contained no detectable fibers. The source of the high fiber burden in S17 is unknown and could not be determined because detailed fiber-source data were not collected.

The universal detection of polyethylene terephthalate in meconium fragments (100% of samples) is consistent with widespread environmental and consumer-product exposure to PET. Polyethylene terephthalate is widely used in packaging and synthetic textiles (Geyer et al., 2017). In Dhaka, where packaged drinking water and plastic food containers are extensively used, environmental polyethylene terephthalate contamination is well documented (Parvin et al., 2022). (Zhang et al., 2021) reported polyethylene terephthalate concentrations in infant feces approximately ten times higher than in adult feces, underscoring the disproportionate burden on neonates and early-life populations. The exclusive identification of nylon as the fiber polymer across all 25 fiber-positive samples is distinctive and may be consistent with exposure to synthetic microfibers from indoor environments, including clothing and household textiles; however, specific exposure sources were not determined. Nylon fibers have been previously reported in human breast milk and placental tissue (Ragusa et al., 2022, 2021).

NaCl-based separation effectively recovers low-density polymers such as polyethylene and polypropylene but may underrepresent denser polymers such as PET (∼1.38 g/cm³) and PVC (∼1.3-1.45 g/cm³). Accordingly, polymer-specific recovery bias toward lower-density particles cannot be excluded. This method was selected for its established use in biological tissue studies and compatibility with downstream spectroscopy (Schwabl et al., 2019).

Together, these results show that MPs were widely detectable in meconium from this cohort and were characterized primarily by polyethylene terephthalate-rich fragments and nylon fibers. Before assessing whether this neonatal particle burden was accompanied by concurrent metal exposure, we examined maternal breast milk as a complementary exposure matrix to determine whether maternal/lactational microplastic profiles paralleled those observed in meconium.

### 3.3 Maternal breast milk microplastics as a complementary exposure context

Microplastics were detected in all analyzed maternal breast milk samples (n = 20). Fragments were the dominant particle type, with a median count of 204 particles/mL (IQR: 145.8-376.5) (Table 1). Fragment sizes were generally smaller than those observed in meconium, with a median size of 60 µm compared with 100 µm in meconium, and 38.8% of breast milk fragments measuring <50 µm (Fig. 1B). This shift toward smaller fragment sizes in breast milk is broadly consistent with previous studies reporting that breast-milk microplastics are often dominated by small particles, although methodological differences can influence the reported size distribution (Adjama et al., 2024; Ragusa et al., 2022). Fibers were detected in 16/20 samples, with a median of 3 fibers/mL. Raman spectroscopy identified polypropylene as the most frequently detected fragment polymer in breast milk (85.0%), followed by polyethylene terephthalate (80.0%), high-density polyethylene (55.0%), polystyrene (45.0%), and low-density polyethylene (15.0%). As in meconium, all fibers were identified as nylon.

Maternal breast milk microplastic counts were compared with neonatal meconium microplastic counts to assess rank-order concordance between the two matrices. The observed negative correlation for fragments (Spearman ρ = -0.462, p = 0.046) and the borderline negative trend for total microplastics (ρ = -0.447, p = 0.055) indicate an absence of positive concordance between maternal breast milk and neonatal meconium particle burdens in this subset. Because the matrices represent different biological compartments and exposure windows, these correlations should not be interpreted as direct measures of placental transfer efficiency. Two ceiling-coded meconium samples (S20 and S21, both recorded at 1000 or more particles per gram) may affect the rank-order correlation estimate, and this finding should be interpreted with appropriate caution.

Polymer concordance within individual mother–neonate pairs was partial. Polyethylene terephthalate was detected in both matrices in 15 of 19 pairs, making it the most commonly shared polymer. However, the degree of polymer overlap varied considerably across pairs: some pairs shared three or four polymer types, while two pairs (S21, S22) shared no fragment polymer types between meconium and breast milk. This within-pair polymer discordance indicates that the two matrices are not interchangeable measures of microplastic exposure and may capture different exposure windows, sources, or biological processes.

Importantly, no sex-based difference was observed in maternal breast milk microplastic counts. Mothers of male neonates had a median total microplastic count of 206 per milliliter compared with 220 per milliliter for mothers of female neonates (Mann-Whitney p = 0.847) (Fig. S6). The absence of a corresponding sex difference in breast milk indicates that the postpartum breast-milk pattern did not parallel the borderline sex difference observed in meconium. However, breast milk collected postpartum cannot distinguish maternal gestational exposure from placental or fetal processes, and the meconium sex association should therefore remain hypothesis-generating.

Direct quantitative comparison of microplastic counts between meconium and breast milk was not performed because the reporting units differ (particles per gram wet weight for meconium versus particles per milliliter for breast milk). The Spearman correlation appropriately tests rank-order agreement regardless of unit differences, and this was the primary analytical approach used for the paired comparison. Overall, breast milk provided a complementary maternal exposure context but was not an interchangeable measure of neonatal meconium MP burden. Additional breast-milk microplastic characterization is provided in Supplementary Materials 1. We therefore next examined whether the neonatal meconium MP burden was accompanied by concurrent metal exposure.

### 3.4 Neonatal meconium metal burden

Returning to the neonatal meconium matrix, all fifteen measured elements were detected in 100% of the 28 analyzable meconium samples (samples 18 and 19 were excluded because of insufficient material). Median concentrations (µg/g dry weight) ranked as follows: Zn (383.3) > Fe (188.0) > Cu (102.9) > Mn (54.9) > Al (15.9) > Cr (3.9) > Se (1.7) > Ni (1.2) > Pb (1.2) > Hg (0.48) > Co (0.21) > As (0.19) > Cd (0.054) > V (0.038) > Be (0.003) (Fig. 2A, B).

**Fig. 2:**
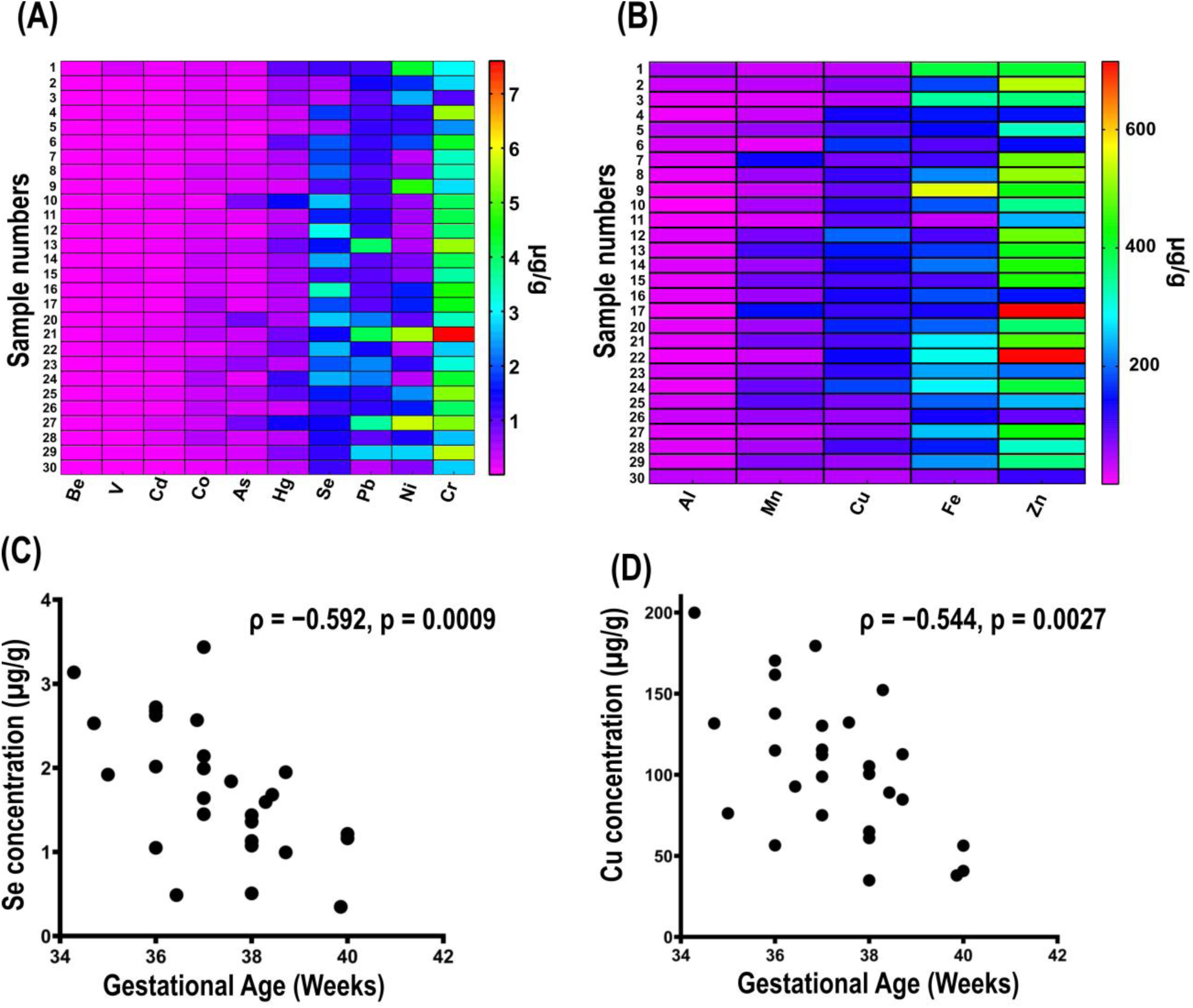
Meconium metal burden and gestational-age associations. **(A)** Heatmap of lower-abundance metals: beryllium, vanadium, cadmium, cobalt, arsenic, mercury, selenium, lead, nickel, and chromium. **(B)** Heatmap of higher-abundance metals: aluminum, manganese, copper, iron, and zinc. Panels were separated by concentration range to improve visual resolution. Concentrations are expressed as µg/g dry weight. **(C)** Spearman correlation between selenium concentration and gestational age. **(D)** Spearman correlation between copper concentration and gestational age. Correlation coefficients and unadjusted p-values are shown in panels C and D; both associations remained significant after Benjamini–Hochberg correction (selenium adjusted *p* = 0.014; copper adjusted *p* = 0.021; *n* = 28).

Concentrations of essential elements were broadly consistent with meconium profiles reported from other populations (Fogarty et al., 2025; Türker et al., 2006).

Among toxicologically relevant metals, lead and chromium were notable. Median Pb concentration was 1.18 µg/g, higher than values reported in New York City meconium but lower than those reported from an industrial Turkish city (Pavilonis et al., 2022; Türker et al., 2006).

Four samples exceeded 2.5 µg/g Pb, which is noteworthy given the absence of a known safe threshold for neonatal lead exposure (Tchounwou et al., 2012). Median Cr concentration was 3.92 µg/g, consistent with concern regarding legacy tannery-associated contamination in Dhaka (Hossain et al., 2021; Tanvir et al., 2022). Mercury was also detectable in all samples, with a median of 0.48 µg/g.

Inter-metal correlation analysis identified 11 nominal associations at an unadjusted Spearman p < 0.05. Nominal positive correlations among toxic metals included As-Cd (ρ = 0.524, p = 0.004), Pb-Cd (ρ = 0.474, p = 0.011), Pb-Cr (ρ = 0.404, p = 0.033), Cd-Hg (ρ = 0.407, p = 0.032), and Cr-V (ρ = 0.431, p = 0.022). Among essential metals, nominal positive correlations included Se-Cu (ρ = 0.775, p < 0.001), Mn-Co (ρ = 0.508, p = 0.006), Be-Fe (ρ = 0.429, p = 0.023), and Ni-Fe (ρ = 0.391, p = 0.040), whereas Ni-Cu (ρ = −0.518, p = 0.005) and Se-Ni (ρ = −0.490, p = 0.008) were inversely correlated. Two samples (S21 and S27) showed concurrent elevation across multiple toxic metals. Overall, these patterns are compatible with co-occurring metal exposures but should be interpreted descriptively because no multiple-testing correction was applied to the pairwise inter-element analysis. Taken together, the universal detection of fifteen metals with nominal inter-element correlations documents a complex multi-element prenatal exposure profile in this urban Dhaka cohort. An extended result and discussion on heavy metal burden have been given in Supplementary Materials 1. Whether this metal burden tracked microplastic burden within individual neonates was examined next.

### 3.5 Microplastic-metal relationships in meconium

To assess whether meconium microplastic burden tracked concurrent metal exposure, Spearman correlations were calculated between total microplastic count and each measured metal using samples with complete paired data (n = 26). Among the fifteen metals examined, only Al showed a nominally significant inverse correlation at the unadjusted level (ρ = -0.404, p = 0.041). However, after Benjamini–Hochberg false-discovery-rate correction, no MP-metal correlations remained significant (all adjusted p > 0.20).

A complementary median-stratified comparison (high versus low microplastic burden; n = 13 per group) yielded a similar pattern. Al concentrations were lower in the high-MP group (median: 13.2 vs. 20.1 µg/g; p = 0.014), and Cd showed a borderline trend toward higher concentrations in the high-MP group (median: 0.066 vs. 0.043 µg/g; p = 0.051), but neither association remained significant after multiple-testing correction.

Taken together, these results do not support a robust monotonic relationship between meconium microplastic burden and meconium metal concentrations in this cohort. Although MPs can adsorb metals under environmental conditions, the biological relevance of such interactions during prenatal exposure remains uncertain. In this context, microplastic particles and metals may arise from partially overlapping or distinct exposure and accumulation processes, although the present data do not resolve these pathways. The use of particle count rather than surface-area-based MP metrics and the need for multiple-testing correction may also reduce sensitivity to weaker associations. The borderline Cd trend may nevertheless warrant follow-up in larger cohorts, given prior evidence that cadmium can sorb to microplastic surfaces under environmental conditions (Wang et al., 2023; Zhou et al., 2020).

### 3.6 Demographic association

Male neonates (n = 17) showed a higher meconium microplastic burden than female neonates (n = 11), with median total MP counts of 194 versus 134 particles/g wet weight, respectively (Mann-Whitney U = 136, p = 0.045) (Fig. S4). Given the unequal group sizes and modest sample size, this borderline difference should be interpreted cautiously and regarded as hypothesis-generating rather than conclusive.

No significant association was observed between gestational age and meconium microplastic burden (Spearman ρ = 0.126, p = 0.523). In contrast, two essential elements showed significant inverse correlations with gestational age. Selenium was negatively correlated with gestational age (ρ = −0.592, p = 0.0009; Benjamini–Hochberg adjusted p = 0.014), as was copper (ρ = −0.544, p = 0.0027; adjusted p = 0.021) (Fig. 2C, D; Fig. S5). These associations may reflect differences in meconium concentration or deposition dynamics across gestation, although this interpretation remains tentative.

No sex-based differences in meconium metal concentrations remained significant after multiple-testing correction. Chromium showed a higher median concentration in males than females (4.04 vs. 2.83 µg/g; p = 0.077), and iron showed a similar trend (208.5 vs. 156.5 µg/g; p = 0.085), but neither reached statistical significance. Because meconium primarily reflects prenatal or near-birth exposure, postnatal variables such as feeding mode were not analyzed as explanatory factors for meconium contaminant burden. Additional demographic analyses and discussion are provided in Supplementary Materials 1. As an additional exploratory dimension, bacterial community profiles were examined in a subset of meconium samples.

### 3.7 Exploratory meconium bacterial profiles

Exploratory genus-level 16S profiling of 12 meconium samples revealed low-complexity but highly heterogeneous bacterial communities (Fig. 3). Individual samples were frequently dominated by one or a few genera, including *Staphylococcus*, *Enterobacter*, *Klebsiella*, *Enterococcus*, *Clostridium*, *Escherichia*, and *Mammaliicoccus*. Male samples showed moderately higher Shannon diversity than female samples (Δmean = 0.302), although the difference remained statistically uncertain (P = 0.084). Sex similarly explained 16.5% of Bray-Curtis community variation (PERMANOVA R² = 0.165, P = 0.084), indicating a suggestive rather than definitive sex-associated pattern.

**Fig. 3:**
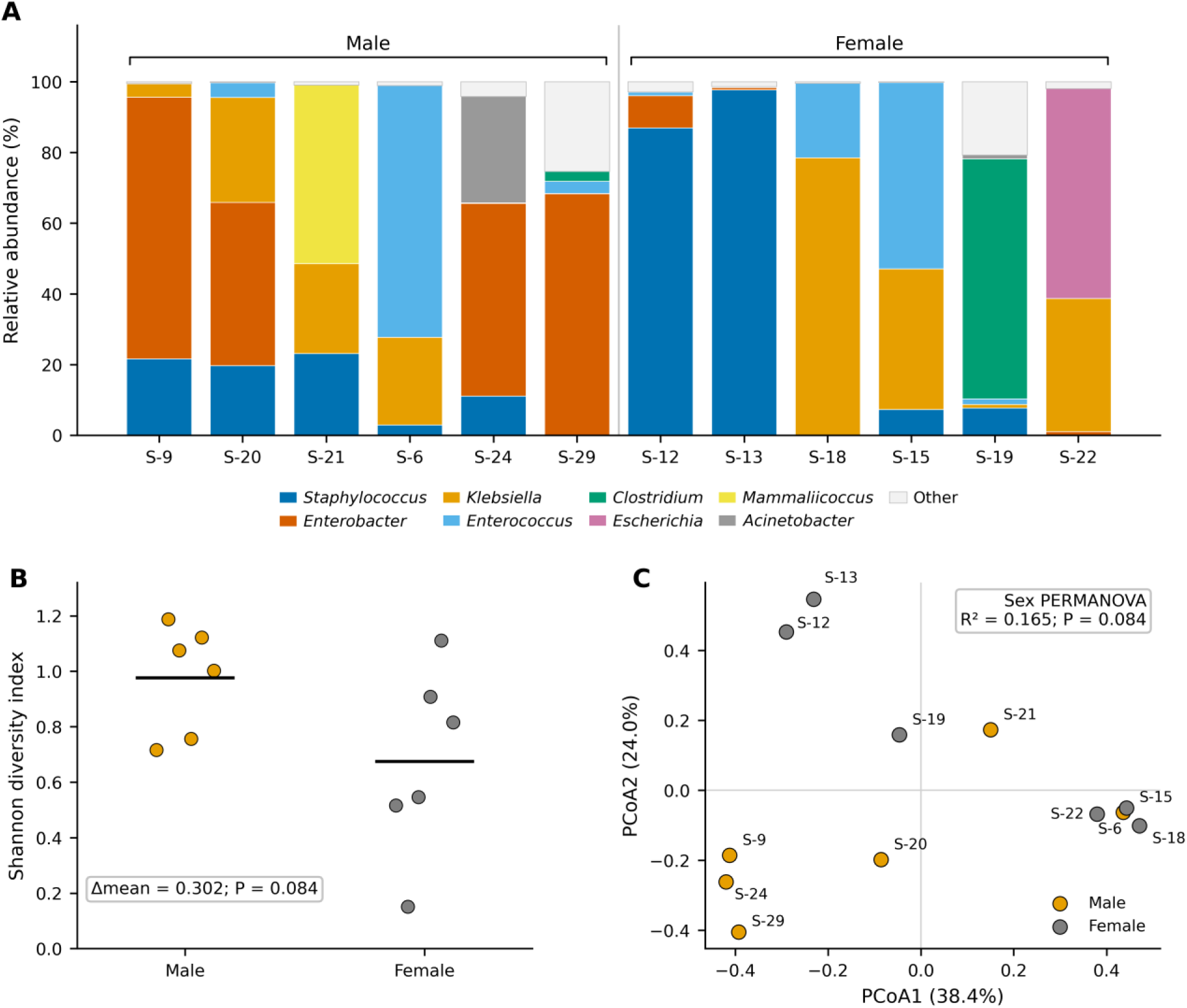
Exploratory genus-level bacterial profiles and community structure in neonatal meconium. **(A)** Genus-level relative abundance profiles for the 12 neonatal meconium samples. The eight most abundant genera across the combined dataset are shown individually, with remaining genera grouped as “Other.” Samples are arranged by neonatal sex for visualization. Profiles were generally low-complexity but highly heterogeneous, with individual samples frequently dominated by different genera, including *Staphylococcus*, *Enterobacter*, *Klebsiella*, *Enterococcus*, *Clostridium*, *Escherichia*, and *Mammaliicoccus*. **(B)** Genus-level Shannon diversity for male and female neonates. Points represent individual samples and horizontal lines indicate group means. Male samples showed moderately higher Shannon diversity on average (male–female Δmean = 0.302), although the difference did not reach conventional statistical significance by exact permutation testing (P = 0.084). **(C)** Principal coordinates analysis of genus-level Bray-Curtis dissimilarities. PCoA1 and PCoA2 explained 38.4% and 24.0% of the variation, respectively. Samples are colored by neonatal sex and labelled by sample identifier. Sex explained 16.5% of overall community variation by PERMANOVA, but the association remained statistically uncertain (R² = 0.165, exact P = 0.084). Together, the panels indicate substantial interindividual heterogeneity in early meconium bacterial profiles, with a suggestive but not definitive sex-associated pattern in diversity and community composition.

Total microplastic burden, fragment abundance, and categorical microplastic burden showed little evidence of association with overall community composition. Fiber abundance, however, was the strongest variable in the exploratory metadata screen (R² = 0.238, P = 0.0034, FDR = 0.0306). The association was stable across transformations but weakened after adjustment for neonatal sex and after exclusion of the two zero-fiber samples (Fig. S8; Tables S11-S12). Fiber abundance should therefore be regarded as a hypothesis-generating correlate rather than evidence of an independent microplastic effect.

The taxon-dominated structure is compatible with recent observations from early infant gut metagenomics. Sinha et al., 2026 reported frequent dominance by one or a few taxa at two weeks of age, including *Escherichia coli* and *Staphylococcus epidermidis*, and identified Cesarean delivery as a major determinant of early community composition. Although their samples were postnatal feces rather than meconium, these findings provide broader context for the low-complexity, heterogeneous profiles observed here. Interpretation of meconium bacterial signals nevertheless remains contentious because of the low-biomass matrix and potential for perinatal acquisition or contamination (Dos Santos et al., 2021; Kennedy et al., 2021; Turunen et al., 2021). Accordingly, these profiles are best viewed as exploratory biological context rather than evidence of a defined prenatal microbiome. Additional analyses are provided in Supplementary Material 1 (Fig. S7-S8; Tables S11-S12).

## 4. Conclusion

This study documents concurrent microplastic and trace-element occurrence in neonatal meconium from an urban South Asian birth cohort. Microplastics were detected in all analyzable samples, with a median burden of 149 particles/g wet weight and profiles dominated by PET fragments and nylon fibers, demonstrating ubiquitous particle detection in this cohort. All fifteen measured elements were detected in analyzable meconium samples. No microplastic–element association remained significant after multiple-testing correction, providing no evidence in this cohort that total particle burden consistently covaried with elemental concentrations. Maternal breast milk showed a partly overlapping but distinct microplastic profile and no positive rank-order concordance with paired neonatal meconium counts, supporting its interpretation as a complementary rather than interchangeable exposure matrix. The nominally higher microplastic burden observed in male neonates and the exploratory bacterial-community patterns require confirmation in larger cohorts. Together, these findings demonstrate the utility of meconium for integrated early-life biomonitoring while highlighting the need for larger studies capable of resolving exposure sources, biological transfer pathways, and potential health relevance.

## Supporting information

Supplementary materials 3 (Raw data)

## Acknowledgments

The authors thank Maksudur Rahman Nayem for support with 16S profiling of the meconium samples.

## Declaration of Generative AI and AI-assisted technologies in the writing process

The author(s) used Grammarly, Claude, and ChatGPT for language editing and ChatGPT, Gemini, and Claude for technical assistance in software setup and data analysis scripting. All content was subsequently reviewed and approved by the author(s), who assume full responsibility for the final manuscript.

## CRediT authorship contribution statement

**Ifthikhar Zaman:** Writing – review & editing, Writing – original draft, Visualization, Validation, Methodology, Investigation, Formal analysis, Data curation, Conceptualization. **Mahdi Muhammad Moosa:** Writing – review & editing, Writing – original draft, Visualization, Validation, Methodology, Investigation, Formal analysis, Data curation, Conceptualization. **Eshra Sultana:** Validation, Methodology, Investigation, Formal analysis. **Rownok Ara Sara:** Validation, Methodology, Investigation, Formal analysis. **Nushrat Jahan:** Validation, Methodology, Investigation, Formal analysis. **Sukanna Mysha:** Validation, Methodology, Investigation, Formal analysis. **Nazifa Tabassum Tasnim:** Validation, Writing – review & editing. **Mohammad Moniruzzaman:** Validation, Methodology, Investigation, Formal analysis, Data curation. **Md Yasir Arafat:** Validation, Methodology, Investigation, Formal analysis, Data curation. **M. Mahboob Hossain:** Writing – review & editing, Validation, Supervision. **Nadia Sultana Deen:** Writing – review & editing, Validation, Supervision.

## Data Availability

The complete raw data are provided as Supplementary Materials 3.

## Ethics Approval

This study was approved by the BRAC University Institutional Review Board (IRB) (Approval no: BRACUIRB120240014).

---

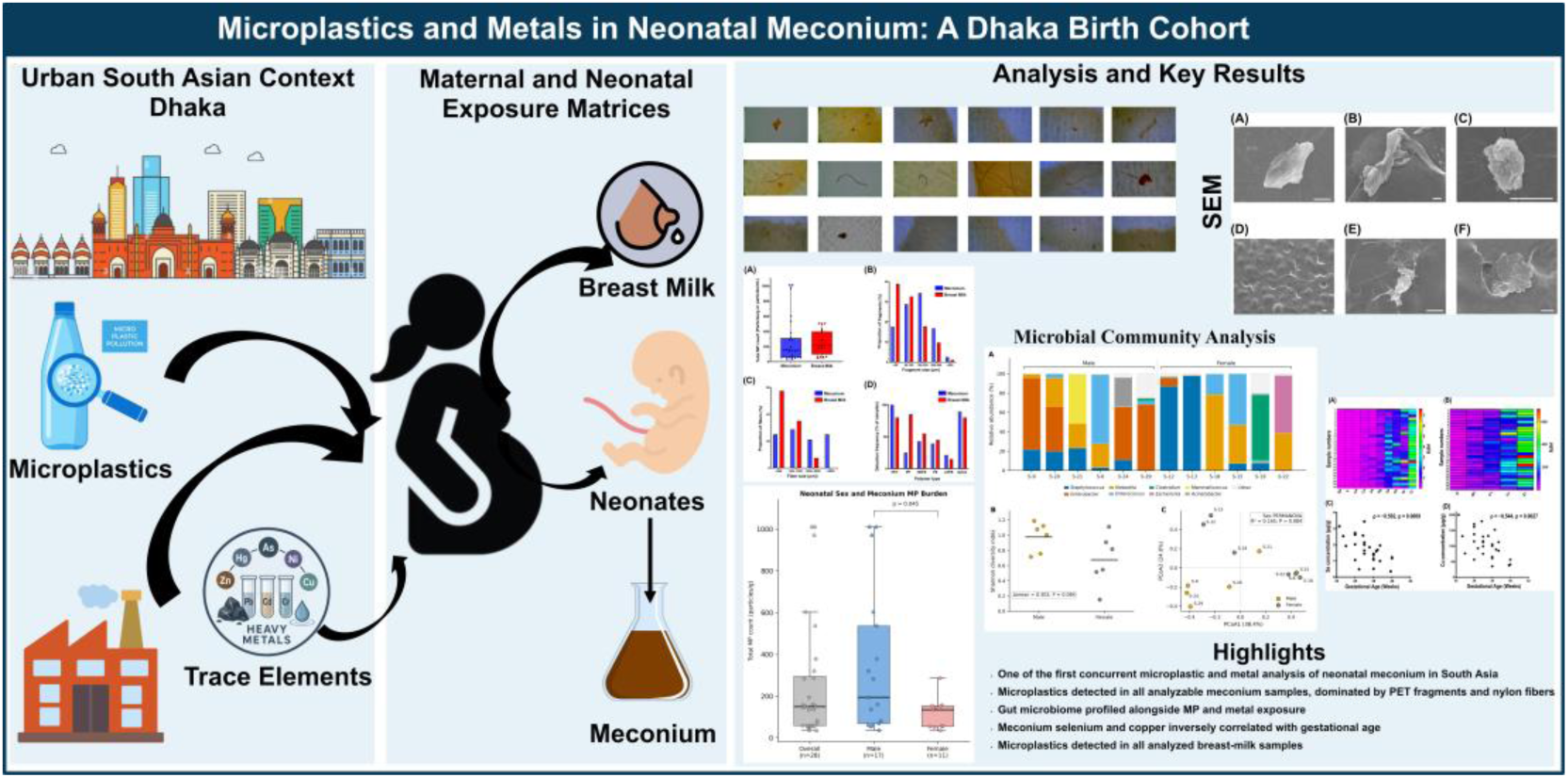
Graphical Abstract.

