## Supplementary materials 1 for "Neonatal meconium reveals concurrent microplastic and metal exposure in an urban South Asian birth cohort"

**S1. Experimental** **Details**

**S1.1. Study Design and Ethical Approval**

This hospital-based, cross-sectional biomonitoring study was conducted at Ad-Din Medical College Hospital between August and October 2025. Thirty neonates were enrolled during the study period. No formal power calculation was performed because the study was exploratory. The study protocol was approved by the BRAC University Institutional Review Board (IRB) (Approval no: BRACUIRB120240014) and conducted in accordance with the Declaration of Helsinki. Written informed consent was obtained from mothers or legal guardians prior to sample collection.

Eligible neonates were those delivered at the study site during the sampling period with a gestational age of ≥34 weeks and no major congenital or gastrointestinal malformations. Neonates born to mothers with clinically diagnosed infectious diseases (e.g., HIV, hepatitis B or C, or systemic infections) were excluded. Cases in which informed consent could not be obtained were also excluded.

**S1.2. Sample Collection**

Meconium samples were collected from 30 neonates within 0–24 h of birth. First-pass meconium was collected by trained nursing staff directly into pre-cleaned sterile glass vials following protocols established for microplastic analysis in neonatal biological matrices (Braun et al., 2021; Liu et al., 2023). Samples were stored at −20 °C until analysis. Two samples yielded insufficient volume for trace-element analysis, and two others were lost during microplastic processing. Thus, 28 samples were available for individual microplastic and trace-element analyses, and 26 for combined analyses.

Breast milk samples (n = 20) were collected from lactating mothers by manual expression into pre-cleaned glass vials following previous studies (Ragusa et al., 2022). Two commercially available infant formula products were obtained from the study hospital. Formula milks were already prepared by the nurses to give to the neonates. This study collected 5 mls of the prepared formula milk and processed for MP analysis identically to the other breast milk samples. Formula inclusion was exploratory, intended to provide preliminary contamination data rather than a representative market survey.

All collection containers were pre-cleaned, rinsed with filtered deionized water, and verified free of microplastic contamination. To minimize airborne and procedural contamination, non-plastic materials (glass and metal) were used throughout sample handling and storage.

**S1.3. Microplastic Isolation and Identification**

Microplastic extraction was performed on meconium (0.5 g per sample), breast milk (1 mL), and infant formula (1 mL). All procedures used glass and metal equipment, following established microplastic protocols. Personnel wore cotton laboratory coats, nitrile gloves, and cotton face masks throughout. Reagents and deionized water were pre-filtered (Whatman® ashless filter paper, Grade 41) to remove microplastic contamination. Glassware was cleaned with laboratory detergent, rinsed with filtered deionized water, autoclaved, and oven-dried.

Procedural blanks (no biological material) were processed alongside each batch. Blank-corrected MP counts were obtained by subtracting the mean blank count for the corresponding analytical batch. Corrected counts ≤ 0 were recorded as zero. During analysis, blanks contained zero MPs (both frags and fiber).

Organic matter was digested using Fenton oxidation. Samples were treated with 30% hydrogen peroxide (H₂O₂) and ferrous iron catalyst (Fe²⁺, as FeSO₄·7H₂O) at 60°C under continuous stirring for 48 h. Additional H₂O₂ was added as needed until the organic material was fully degraded. Fenton's reagent was selected for its validated efficiency in digesting organic-rich biological matrices while preserving common polymer types (Hurley et al., 2018; Tagg et al., 2016). Density separation was then performed using saturated sodium chloride (NaCl) solution (~1.2 g/cm³). Samples were settled for 48 h, and the supernatant was collected. NaCl-based separation effectively recovers low-density polymers such as polyethylene and polypropylene but may underrepresent denser polymers such as PET (~1.38 g/cm³) and PVC (~1.3–1.45 g/cm³). Accordingly, polymer-specific recovery bias toward lower-density particles cannot be excluded. This method was selected for its established use in biological tissue studies and compatibility with downstream spectroscopy (Schwabl et al., 2019).

The supernatant was vacuum-filtered through glass microfiber filters (0.10 µm, JVLAB), dried in a desiccator, and examined under a stereomicroscope (SOPTOP SZN71; 40× magnification). Particles ≤5 mm were enumerated and classified by morphology (fragment or fiber) and approximate size.

Polymer identification of a subset of frags and fibers was performed using macro-Raman spectroscopy (HORIBA MacroRAM). Spectra were acquired over 400–1800 cm⁻¹ and matched against reference libraries via the OpenSpecy platform (Cowger et al., 2021). Assignments were accepted at a hit quality index (HQI) threshold of ≥45%. Spectra below this threshold were classified as unidentified. The subset was chosen on the basis of morphology, identified during stereomicroscopy. From each sample, more than 8 frags and fibers were selected. Selected particles were further examined by scanning electron microscopy (SEM; JEOL JSM-7610F) to confirm surface morphology.

**S1.4. Trace-Element Analysis**

Fifteen trace elements (As, Se, Pb, Cd, Cr, Ni, Cu, Zn, Hg, Mn, Co, Al, Be, V, and Fe) were quantified in meconium by inductively coupled plasma mass spectrometry (ICP-MS; PerkinElmer NexION 2000), following established protocols for elemental analysis in neonatal biological matrices (Fogarty et al., 2026; Michelsen-Correa et al., 2021).

**Sample Analysis:** Meconium samples were digested using microwave-assisted acid digestion adapted from US EPA Method 3051A. Approximately 0.1-0.5 g of dried, homogenized meconium was placed in tetrafluoromethoxyl (TFM) vessels. Under a safety shield, 7 mL of 65% ultra-pure nitric acid (HNO₃) and 3 mL of 30% hydrogen peroxide (H₂O₂) were added (HNO₃:H₂O₂ = 7:3, v/v) following microwave digestion application note HPR-BIO. Vessels were sealed and digested at 200 °C for 30 min using a Milestone Start D microwave digestion system (Milestone, Italy). After cooling to room temperature, the digested solution was extracted with deionized water, transferred to a 25 mL volumetric flask, filtered, and stored for ICP-MS analysis. The nebulized sample can enter (~0.85 ml/ min) the core of the argon (Ar) plasma of the ICP-MS system. A quadrupole mass detector determined the trace metal ions generated at high-temperature plasma as mass to charge ratio (*m*/*z*). An ICP-multi-element standard solution XIII (Merck, Germany) was used to prepare a standard calibration curve at ppb levels. An appropriate amount of the internal standard mixture (45Sc, 73Ge, 89Y, 115In, 185Re, 193Ir, and 209Bi) was then added before being characterized by ICP-MS to reduce possible sources of error, such as instrumental drift, matrix effects, and variations in sample introduction. We performed reagent blanks, filter blanks, spike samples, and recoveries of the SRM sample in order to ensure accuracy and precision with minimal analytical errors. The details are as follows.

**Internal Standard:** Stringent quality assurance measures were implemented in this study to ensure the reliability and accuracy of the obtained results. At the core of these initiatives was the deliberate utilization of internal standards in Inductively Coupled Plasma Mass Spectrometry (ICP-MS) analysis. Selected internal standards, consisting of known concentrations of non-analyte elements, were carefully chosen and added to standard solutions and sample matrices before analysis. The main goal of using internal standards was to reduce possible sources of error, such as instrumental drift, matrix effects, and variations in sample introduction. This would improve the accuracy and precision of quantitative measurements of the samples. We closely monitored the stability of the instrument throughout the analytical runs by observing the response of the internal standard in relation to the target analytes. Any variations in the internal standard signal were used to identify instrumental drift, which led to prompt corrective actions such as recalibration or adjustment of instrument parameters. In addition, the use of internal standards helped to address issues caused by variations in the samples or interferences, resulting in accurate measurement of the desired analytes. Therefore, internal standards play a crucial role in ensuring accurate comparisons of analyte concentrations and reliable interpretation of results by standardizing variations in analytical responses across different sample matrices. Regular evaluations were conducted to assess the performance of the analytical instrument, focusing on the consistency and reproducibility of internal standard signals. Thorough investigations were conducted whenever deviations or inconsistencies were observed in internal standard responses. These help to identify potential sources of error, such as instrument malfunctions or sample preparation artifacts. An internal standard also has a significant impact on data validation and quality control. Their methodical integration into the analytical workflow offered a strong framework for evaluating method performance, detecting possible outliers, and guaranteeing the reliability of the generated data. In summary, the deliberate use of internal standards within the quality assurance protocol demonstrated the dedication to producing accurate and dependable data, thereby enhancing the trustworthiness and credibility of the study results.

**Quality assurance and quality control:** All glassware used in this experiment was cleaned and rinsed with ultrapure acid and water to reduce the possibility of contamination during sample preparation and analytical operations. The performance of the ICPMS equipment was checked by the NexION Setup solution (PerkinElmer, USA) before calibration with standards. A five-point calibration curve, R^2^ *>* 0.9995, was achieved over a dynamic range of 1.0 - 100 ppb. A minimum of three replicates was conducted to confirm instrument stability. We considered the maximum relative standard deviation (RSD) of 5-8%. The instrument’s detection (DL) and quantification (QL) limits were defined when the response for a specific metal was three and ten times greater than the background noise, respectively. A standard reference material, SRM NIST 1648a, from the National Institute of Standards (NIST) was used to check the recovery percentage of elements (As, Al, Cd, Cr, Cu, Ca, Fe, Pb, Mg, Mn, Ni, K, Na, Ti, V, Zn) in the sample. Element recoveries obtained using SRM NIST 1648a are presented in Table S1. The Reagent blanks, filter blanks, spike sample, and recoveries of SRM samples were performed as part of the quality control process to minimize error. Corrections to the trace metal concentrations were made based on recovery and analysis of blank samples.

**S1.5. Exploratory Meconium Microbial Community Analysis**

**Community profiling and diversity:** Emu abundance profiles were collapsed to genus level and analyzed as relative abundances. Shannon diversity was calculated from the complete genus-level profile, while between-sample community structure was assessed using Bray-Curtis dissimilarity and principal coordinates analysis (PCoA), consistent with previous analyses of full-length Oxford Nanopore 16S datasets processed using Emu (Yeo et al., 2024).

**Metadata association analyses:** Exploratory associations were evaluated for neonatal sex, gestational age, term status, feeding, baby-product category, MP category, and continuous total MP, fragment, and fiber abundances. Binary comparisons of Shannon diversity used exact permutation tests of group mean differences, with Cliff's delta reported as an additional effect-size measure (Cliff, 1993); continuous associations were assessed using Spearman correlation with 9,999 permutations. Total MP and fragment abundance were log10-transformed, whereas fiber abundance was analyzed as log10(1 + fibers/g) to retain zero values. Community-level associations were tested by PERMANOVA (Anderson, 2001), with multivariate dispersion assessed for binary predictors (Anderson, 2006). Binary PERMANOVA comparisons used exact permutations where feasible, whereas continuous predictors were evaluated using 9,999 permutations. Benjamini–Hochberg false-discovery-rate correction was applied separately to the Shannon and PERMANOVA metadata screens (Benjamini and Hochberg, 1995). Stability of effect estimates was assessed by sequential leave-one-out analysis.

**Fiber sensitivity analyses:** Because fiber abundance produced the strongest exploratory community-level association, targeted sensitivity analyses evaluated raw, square-root-transformed, and log10(1 + x)-transformed fiber abundance and adjustment for neonatal sex, fragment abundance, and sequencing run. Adjusted effects were estimated relative to the corresponding reduced models and reported as ΔR² and partial R². Permutation P values were obtained using residual permutation of the fiber predictor after accounting for included covariates, consistent with approaches developed for covariate-adjusted microbiome community testing (Tang et al., 2016). Additional analyses excluded the two samples in which no fibers were detected. Sequencing run was treated solely as a technical sensitivity variable.

**Sequencing-depth robustness:** Genus-level Shannon diversity was evaluated across simulated subsampling depths of 100–61,899 reads, with 100 multinomial resampling iterations performed at each depth using the Emu-estimated genus distribution for each sample. Mean Shannon values and 95% simulation intervals were examined for stabilization with increasing depth. Because Emu provides estimated abundance profiles rather than conventional ASV or OTU count tables, this procedure was used as a sequencing-depth robustness assessment rather than strict count-based rarefaction.

**Computational environment:** Downstream statistical analyses were performed using custom scripts in Python 3.10.20 with NumPy 2.2.6, pandas 2.3.3, and SciPy 1.15.2; graphical outputs were generated using Matplotlib 3.9.1.

**S1.6. Statistical Analysis**

All non-microbiome statistical analyses were performed in Python (v3.14) using the SciPy (v1.17.1) and pandas (v3.0.2) libraries. The complete analysis script (meconium_stats_complete.py) and raw output (supplementary_statistical_output.txt) are provided as supplementary files. All results reported in the manuscript can be reproduced by running the script on the raw data files.

**Normality assessment:** The Shapiro–Wilk test was applied to all continuous variables. The majority of distributions deviated significantly from normality; therefore, non-parametric methods were used throughout. Descriptive statistics are reported as median and interquartile range (IQR; 25th–75th percentile) calculated using pandas' default linear interpolation method.

**Microplastic count normalization:** Meconium MP counts were normalized from the 0.5 g extraction weight to 1 g wet weight by multiplying raw counts by 2. Breast milk and infant formula counts are reported per milliliter (no normalization required). Samples exceeding the counting threshold of 500 fragments per 0.5 g were conservatively recorded as 500 for all statistical calculations. After normalization to 1 g, these correspond to ≥1000 fragments/g. All mean and median values involving these samples are therefore minimum estimates.

Inter-metal correlations: Pairwise Spearman's rank correlations were computed for all 105 metal–metal pairs (15 metals, n = 28 samples). Correlations with p < 0.05 are reported. No multiple testing correction was applied to inter-metal correlations, as these were descriptive rather than hypothesis-testing.

**MP-heavy metal associations:** Two complementary approaches were used on the 26 samples with complete data for both MP and heavy metal analyses (samples 8 and 27 excluded due to meconium loss; samples 18 and 19 excluded due to insufficient volume for heavy metal analysis):

(i) Spearman's rank correlations between total MP count per gram and each of the 15 metals individually. A parallel analysis using fragment count per gram was also performed. Raw p-values were corrected for multiple comparisons using the Benjamini–Hochberg false discovery rate (BH-FDR) procedure across 15 tests.

(ii) Neonates were stratified into high (≥ median) and low (< median) MP burden groups based on the median total MP count per gram (149 particles/g; n = 13 per group). Differences in heavy metal concentrations between groups were assessed using two-sided Mann–Whitney U tests, with BH-FDR correction applied across 15 comparisons.

**Paired mother-neonate MP comparison:** Nineteen valid mother–neonate pairs were identified (one pair was excluded due to meconium loss during processing). Spearman's rank correlation assessed rank-order agreement between meconium MP counts (particles/g wet weight) and corresponding maternal breast milk MP counts (particles/mL). Mann-Whitney U tests compared breast milk MP counts between mothers stratified by neonatal meconium MP burden (high vs. low, based on the median).

**Neonatal sex and MP burden:** Differences in meconium MP counts (total, fragment, and fiber) between male (n = 17) and female (n = 11) neonates were assessed using two-sided Mann–Whitney U tests.

**Neonatal sex and heavy metal concentrations:** Differences between male (n = 19) and female (n = 9) neonates were assessed using two-sided Mann–Whitney U tests for each of the 15 metals (n = 28 samples with heavy metal data). Raw p-values were corrected using BH-FDR across 15 comparisons. Note: this analysis used questionnaire and heavy metal data only (n = 28), without requiring MP data, and therefore includes samples 8 and 27, which lacked MP data but had valid heavy metal measurements.

**Gestational age and MP burden:** Spearman's rank correlations were computed between gestational age (weeks) and meconium MP counts (total, fragment, and fiber; n = 28). Additionally, Mann–Whitney U tests compared MP counts between term (≥37 weeks; n = 17) and late preterm (<37 weeks; n = 11) neonates.

**Gestational age and heavy metal concentrations:** Spearman's rank correlations were computed between gestational age and each of the 15 metals (n = 28 samples with heavy metal data). Raw p-values were corrected using BH-FDR across 15 comparisons.

**BH-FDR correction procedure:** The Benjamini–Hochberg procedure was implemented manually as follows: (1) the 15 raw p-values were ranked from smallest to largest; (2) each p-value was adjusted as: p_adjusted = p_raw × 15 / rank; (3) monotonicity was enforced by a backward pass ensuring that each adjusted p-value was no larger than the adjusted p-value of the next-higher rank; (4) any adjusted p-value exceeding 1.0 was set to 1.0. An adjusted p-value < 0.05 was considered statistically significant.

**Significance threshold:** A two-tailed p-value of < 0.05 was considered statistically significant for all analyses, unless otherwise stated. For analyses involving multiple comparisons across the 15-metal panel, only BH-FDR-corrected p-values < 0.05 were considered significant.

**S2. Detailed Results and Discussion**

**S2.1 Extended breast milk microplastic characterization**

The main text reports that all 20 breast milk samples contained microplastics, with polypropylene as the most frequently detected fragment polymer (85.0%) followed by polyethylene terephthalate (80.0%). This section provides additional detail on particle size distributions and cross-matrix polymer comparisons.

Breast milk fragment sizes (n = 258 measured particles) had a mean of 87.6 ± 87.6 micrometers (median: 60 micrometers; range: 10–550 micrometers), with 38.8% of fragments below 50 micrometers compared with only 17.6% in meconium. This shift toward smaller fragments in breast milk may reflect differences in exposure window, matrix biology, or analytical recovery; the present study cannot distinguish among these possibilities. Fibers in breast milk (n = 40) had a mean length of 503.0 ± 290.3 micrometers (median: 430 micrometers), compared with 920 micrometers median in meconeum. This difference may partly reflect the distinct temporal windows represented by the two matrices, but it does not establish a specific biological transfer mechanism.

The polymer profile shifted notably between matrices. Whereas polyethylene terephthalate dominated meconium fragments at 100% detection, polypropylene was the most frequently detected polymer in breast milk (85.0%), with polyethylene terephthalate at 80.0%. This shift may reflect differences in exposure windows, environmental sources, matrix biology, or analytical recovery; specific sources of PET or PP exposure cannot be assigned from these data. The co-dominance of both polyethylene terephthalate and polypropylene in breast milk, compared with the clear polyethylene terephthalate dominance in meconium, further indicates that the two matrices captured non-equivalent microplastic profiles.

**S2.2 Infant formula microplastic data**

Both commercially available infant formula products analyzed (n = 2) contained 500 or more fragments per milliliter, with all five polymer types detected (polyethylene terephthalate, polypropylene, high-density polyethylene, polystyrene, and low-density polyethylene). Formula I additionally contained 5 fibers per milliliter (nylon), while Formula II contained none. Fragment sizes were very small (10–30 micrometers), substantially smaller than fragments in either meconium (median: 100 micrometers) or breast milk (median: 60 micrometers).

Although the sample size (n = 2) precludes generalization, these findings are consistent with previous reports documenting substantial microplastic contamination in infant formula products. (Li et al., 2020) demonstrated that the sterilization of polypropylene feeding bottles releases millions of microplastic particles into the prepared formula, and (Kadac-Czapska et al., 2024) detected microplastic contamination across multiple commercially available infant formula brands. The detection of all five polymer types in both formula products, compared with the more selective polymer profiles in meconium and breast milk, may reflect the diverse polymer sources in manufacturing, packaging, and preparation equipment. These preliminary observations suggest that formula-fed infants may face additional microplastic exposure through this route, although larger product surveys are needed to establish contamination ranges across brands and preparation methods.

**S2.3 Extended heavy metal discussion**

The main text reports the detection of all fifteen metals in all 28 analyzable meconium samples, with lead, chromium, and mercury at concentrations consistent with intermediate urban exposure. This section provides additional discussion of selected elements, inter-element correlations, and geographic context.

Mercury concentrations (median: 0.48 micrograms per gram dry weight) in this cohort may relate to multiple exposure sources within the Dhaka urban environment. Mercury-containing skin-lightening products are one plausible maternal exposure route, as such products have been shown to contain mercury compounds in systematic assessments of consumer cosmetics (Bastiansz et al., 2022). and dietary sources including rice and fish from mercury-contaminated waterways (Nargis et al., 2020). The Cd-Hg correlation (rho = 0.407, unadjusted p = 0.032) is compatible with, but does not establish, shared exposure sources, as both cadmium and mercury can co-occur in industrial effluents, electronic waste, and certain consumer products.

Chromium concentrations (median: 3.92 micrograms per gram) were notably elevated in this cohort, consistent with Dhaka's well-documented legacy of tannery-associated contamination. The Hazaribagh district historically housed over 200 tanneries that discharged chromium-laden effluent directly into surrounding waterways and soil for decades before partial relocation to the Savar Tannery Industrial Estate (Hossain et al., 2021). Children in industrialized areas of Dhaka have been shown to have elevated urinary chromium compared with less industrialized reference populations (Tanvir et al., 2022). The persistence of elevated chromium in neonatal meconium, even as tannery operations have been partially relocated, is compatible with continuing exposure within a region affected by substantial historical and ongoing environmental metal contamination, although the present study cannot attribute individual neonatal concentrations to specific sources.

The positive correlations among toxic metals (As–Cd, rho = 0.524; Pb–Cd, rho = 0.474; Pb–Cr, rho = 0.404; Cd–Hg, rho = 0.407; Cr–V, rho = 0.431) showed co-variation within the cohort. Because all 105 element–element pairs were evaluated without multiple-testing correction, these correlations are interpreted descriptively and do not establish common-source exposure. The strong Se–Cu correlation (rho = 0.775, p < 0.001) may reflect shared nutritional pathways, as both selenium and copper are essential elements actively and concurrently transported across the placenta via specific transporter proteins to support fetal development (Michelsen-Correa et al., 2021). The inverse Ni–Cu (rho = −0.518) and Se–Ni (rho = −0.490) correlations may suggest competitive absorption at the intestinal level or differential placental transfer kinetics between these element pairs, though mechanistic interpretation is limited at this sample size and these associations were not adjusted for multiple comparisons.

Two individual samples stood out for concurrent elevation across multiple toxic metals. S21, from the Tejgaon area, showed high lead (4.08 micrograms per gram), cadmium (0.21), chromium (7.60), and nickel (5.55 micrograms per gram), while S27, from the Green Road area, showed elevated lead (3.60), arsenic (0.82), cadmium (0.20), mercury (1.41), and nickel (5.79 micrograms per gram). These individuals represent observations of comparatively high co-occurring metal burden, though detailed individual exposure histories were not available. Such individual-level co-exposure clustering has been observed in mining communities in the United States (McDermott et al., 2020) and underscores the importance of assessing cumulative metal mixture exposure rather than individual elements in isolation (Pavilonis et al., 2022). The geographic map of sampling locations (Fig. S2) did not reveal clear spatial clustering of elevated metal concentrations, although interpretation was constrained by the limited spatial resolution of address-level residential data and the small sample size.

**S2.4 Extended demographic associations**

The main text reports a borderline sex difference in meconium microplastic burden (males higher, p = 0.045) and significant inverse correlations between gestational age and selenium and copper concentrations. This section provides an extended analysis and interpretation of these associations.

Regarding the gestational age and microplastic burden, the comparison between term (37 weeks or more; n = 17) and late preterm (less than 37 weeks; n = 11) groups showed no significant difference in total microplastic counts (median 156 versus 82 particles per gram; Mann–Whitney U = 114, p = 0.335) (Fig. S4). The narrow gestational age range in this cohort (34.3–40.0 weeks) and the modest sample size limit the power to detect gestational age-dependent effects on microplastic accumulation. The absence of a strong gestational age–microplastic relationship may also reflect opposing factors: longer gestation allows more time for microplastic accumulation via amniotic fluid ingestion, which would predict a positive correlation, whereas late preterm neonates may have experienced different patterns of placental maturation or amniotic fluid dynamics that could influence particle transfer in the opposite direction.

The inverse selenium–gestational age (rho = −0.592, adjusted p = 0.014) and copper–gestational age (rho = −0.544, adjusted p = 0.021) relationships indicate that neonates born at earlier gestational ages had higher meconium concentrations of these essential elements (Fig. 2C, D). Both selenium and copper are essential trace elements actively transported across the placenta to support fetal development, with selenium required for selenoprotein synthesis and antioxidant defense, and copper required for metalloenzyme function and connective tissue formation (Michelsen-Correa et al., 2021). The higher concentrations at lower gestational ages may reflect relative concentration effects: the same amount of transplacentally delivered element is distributed in a smaller mass of meconium produced over a shorter gestational period, rather than greater absolute maternal exposure or placental transfer. Gestational age-dependent variation in placental transporter expression has been documented, with certain transporters showing higher activity during earlier stages of gestation (Walker et al., 2017).

Regarding sex and heavy metal concentrations, no significant differences were observed between male (n = 19) and female (n = 9) neonates for any of the 15 elements after Benjamini–Hochberg correction (Fig. S5). Chromium trended higher in males (median 4.04 versus 2.83 micrograms per gram; p = 0.077), and iron showed a similar pattern (median 208.5 versus 156.5 micrograms per gram; p = 0.085), but neither reached statistical significance. The unequal group sizes (19 versus 9) and the need to correct for 15 simultaneous comparisons substantially limit power to detect sex-based metal differences. These borderline trends may warrant follow-up in larger, sex-balanced cohorts, particularly given that sex-specific differences in placental function and transporter expression are well documented for essential metals (Meakin et al., 2021; Walker et al., 2017).

Sex-specific differences in placental function provide a possible biological context for the borderline microplastic finding (Meakin et al., 2021; Walker et al., 2017). However, placental microplastic transport was not measured in this study. No sex-based difference was observed in maternal breast milk microplastic counts (male-neonate mothers: median 206 particles/mL; female-neonate mothers: median 220 particles/mL; p = 0.847) (Fig. S6), indicating that the postpartum breast-milk pattern did not parallel the meconium sex pattern. These data cannot distinguish differential gestational exposure from sex-associated placental or fetal processes.

Because meconium primarily reflects prenatal or near-birth exposure, postnatal variables recorded in the questionnaire, including feeding mode, formula brand, and type of infant care product, were not tested against meconium contaminant levels. These postnatal factors were not considered appropriate explanatory variables for contaminant burdens accumulated predominantly during the prenatal period. This analytical decision is consistent with the biological formation timeline of meconium and avoids spurious associations between prenatal biomarkers and postnatal behaviors.

**S2.5 Extended exploratory meconium bacterial profiles**

Genus-level bacterial profiles were determined for 12 neonatal meconium samples. The subset included six male and six female neonates, with three high- and three low-microplastic samples within each sex. Profiles remained low-complexity and highly heterogeneous, with individual samples frequently dominated by one or a few genera (Fig. 3).

The suggestive sex-associated patterns described in the main text were reasonably stable to individual-sample omission. Male samples showed higher mean Shannon diversity than female samples (Δmean = 0.302; Cliff's δ = 0.556; P = 0.084), with leave-one-out differences remaining positive throughout (median = 0.289; range = 0.197–0.389). Sex similarly explained 16.5% of Bray–Curtis variation (R² = 0.165; P = 0.084), with a leave-one-out median R² of 0.173. Female samples showed numerically greater dispersion around their group centroid, but the dispersion difference was not statistically supported (P = 0.126; Table S11).

Total particle abundance, fragment abundance, and categorical microplastic burden showed little evidence of association with overall community composition, explaining 6.8%, 7.0%, and 6.7% of Bray–Curtis variation, respectively (all P > 0.59). Total particle and fragment abundance also showed positive but non-significant associations with Shannon diversity that did not survive false-discovery-rate correction (Table S11).

Fiber abundance produced the strongest exploratory community-composition signal. Log10(1 + fiber abundance) explained 23.8% of Bray–Curtis variation (P = 0.0034; FDR = 0.0306), with a leave-one-out median R² of 0.245. Similar effect estimates were obtained using raw and square-root-transformed fiber abundance and after adjustment for fragment abundance or sequencing run (Fig. S8; Table S12). Adjustment for neonatal sex weakened the association (ΔR² = 0.157; partial R² = 0.188; P = 0.077). Excluding the two zero-fiber samples likewise reduced statistical support despite a remaining moderate effect size (R² = 0.168; P = 0.139). The fiber signal therefore warrants follow-up but cannot be considered independent of sex or the leverage contributed by the zero-fiber observations.

Subsampling-based sequencing-depth analysis showed that genus-level Shannon estimates stabilized rapidly across increasing simulated read depths (Fig. S7), indicating that the observed diversity differences were unlikely to be driven primarily by inadequate sequencing depth. Because Emu provides estimated abundance profiles rather than conventional ASV/OTU count tables, this analysis was used as a sequencing-depth robustness assessment rather than strict rarefaction.

The broader biological context of these taxon-dominated profiles, including comparison with recent early-infant metagenomic data and the continuing uncertainty surrounding bacterial signals in low-biomass meconium, is discussed in the main text. Given the small sample size and limited covariate independence, the analyses presented here remain hypothesis-generating.

**Supplementary Figures**

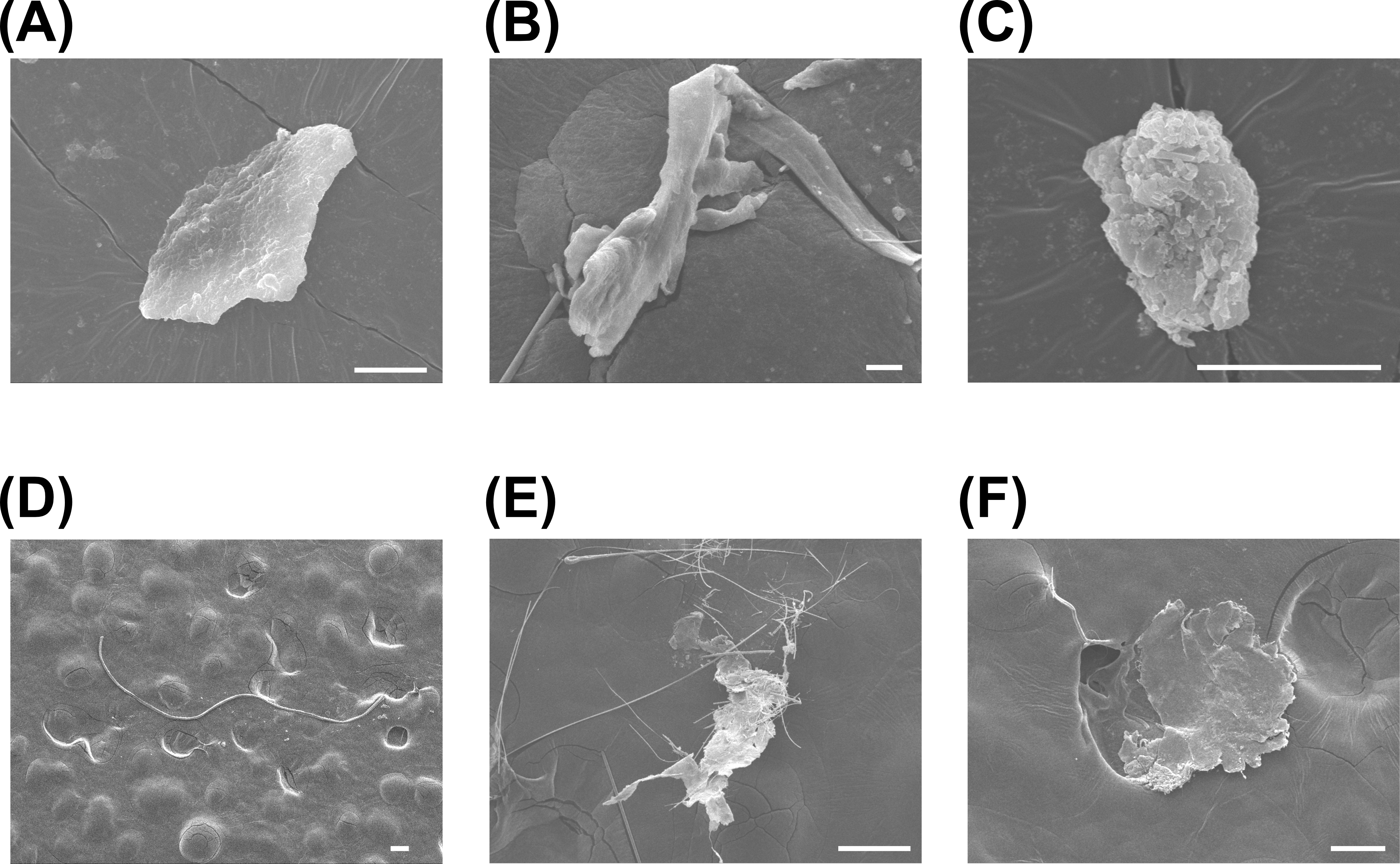

**Fig. S1:** Representative SEM images of particles isolated from neonatal meconium. Panels A-C and E-F show irregular fragment-type particles, whereas panel D shows a fiber. Fiber-type particles were identified as nylon (polyamide) by Raman spectroscopy. Scale bars: 10 µm (A-C) and 100 µm (D-F).

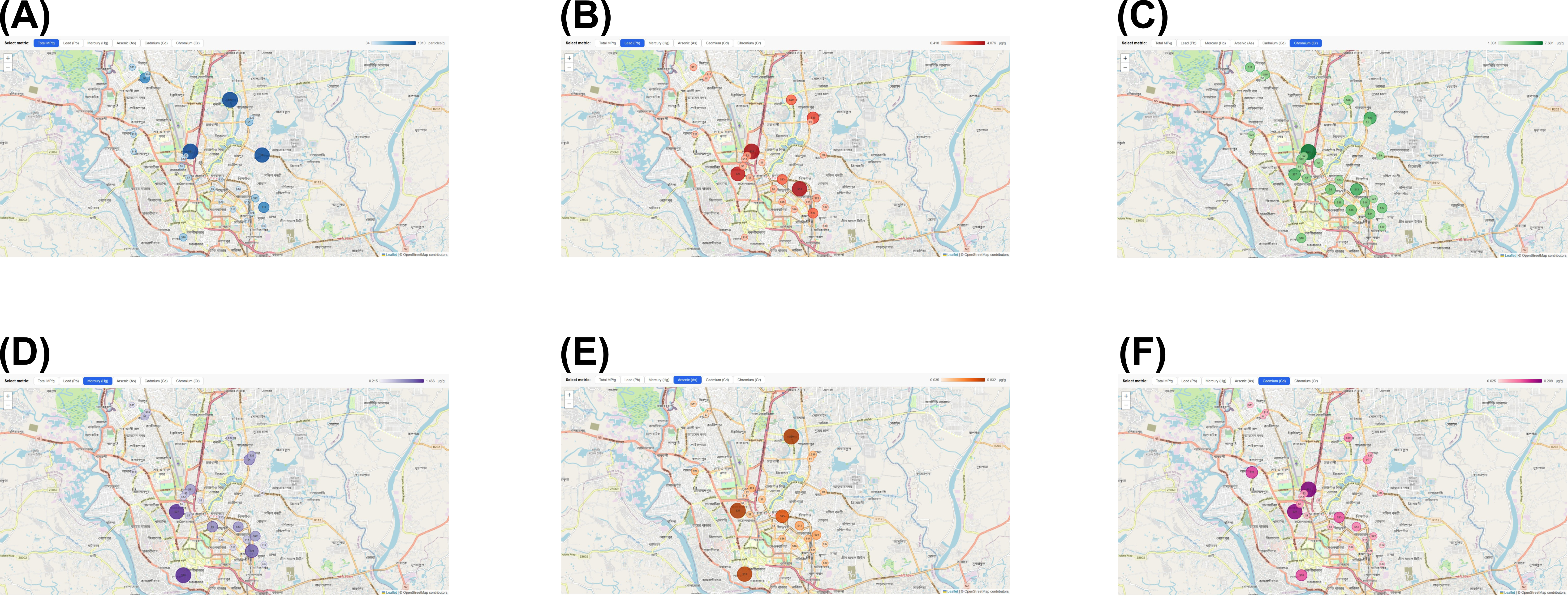

**Fig S2.** Geographic distribution of microplastic and toxic metal concentrations across neonatal meconium sampling locations in Dhaka, Bangladesh. **(A)** Total microplastic count (particles/g wet weight). **(B)** Lead (Pb; µg/g dry weight). **(C)** Chromium (Cr; µg/g dry weight). **(D)** Mercury (Hg; µg/g dry weight). **(E)** Arsenic (As; µg/g dry weight). **(F)** Cadmium (Cd; µg/g dry weight). Bubble size and color intensity are proportional to concentration within each panel. Sample locations (n = 30) are based on maternal residential addresses reported at enrollment; four samples with non-specific "Dhaka" addresses were positioned near the city center with minor spatial offsets for visibility. Dashed circles indicate samples with unavailable data for the selected metric (samples 8 and 27 for MP; samples 18 and 19 for metals). No clear geographic clustering was observed for any metric, although spatial inference was likely limited by the small sample size and the spatial precision of the available residential information. We have added an HTML file of this map for better viewing.

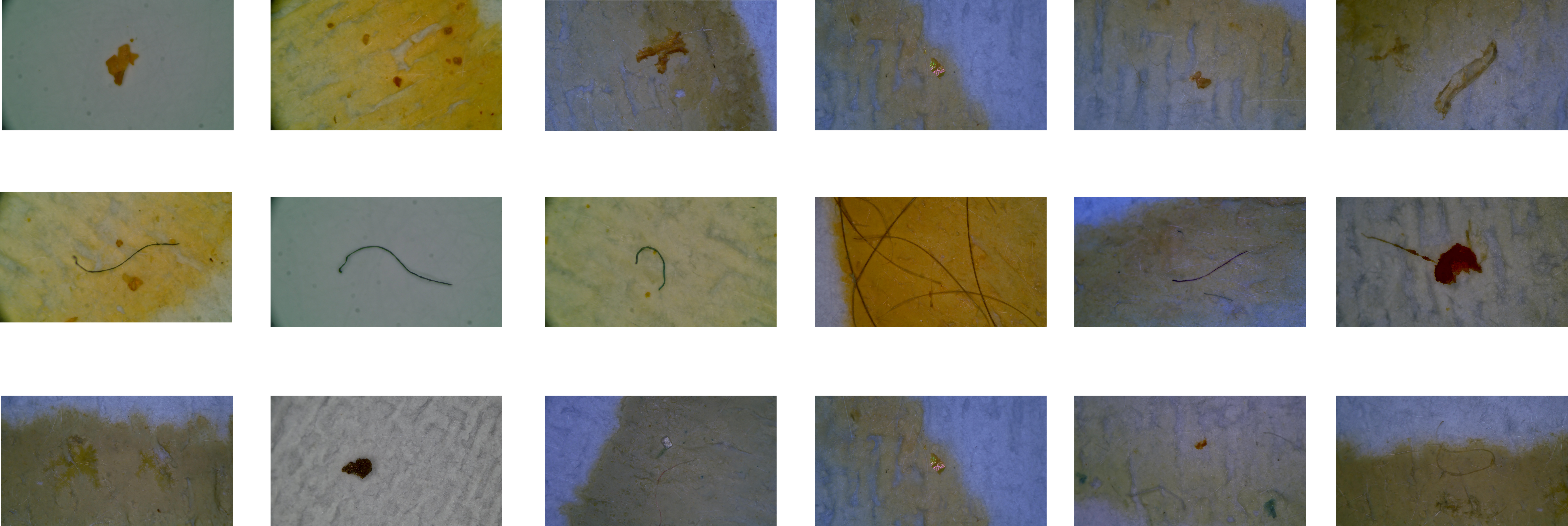

**Fig S3:** Different types of microplastics (fragments and fibers) seen under a stereomicroscope at 40x magnification

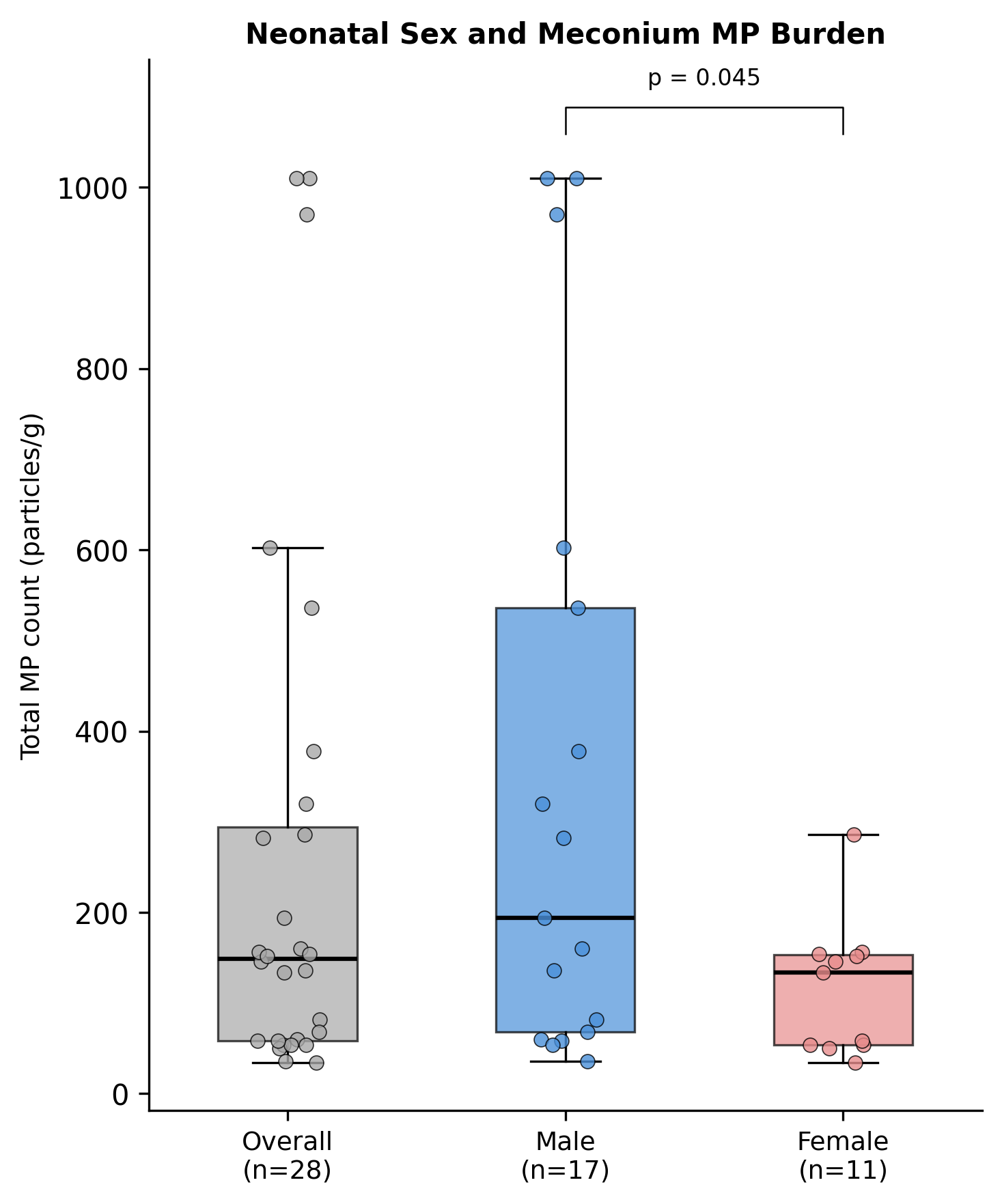

**Fig S4.** Total meconium microplastic counts (particles/g wet weight) by neonatal sex, presented as box-and-whisker plots (median, IQR, min–max) with individual data points. Overall (n = 28; gray), male (n = 17; blue), and female (n = 11; pink) distributions are shown. Male neonates had higher median total MP counts (median: 194 vs. 134 particles/g; Mann-Whitney U = 136, p = 0.045). The borderline significance and unequal group sizes (17 vs. 11) warrant cautious interpretation; this finding is considered hypothesis-generating.

**Fig S5.** Heavy metal concentrations (µg/g dry weight) in neonatal meconium by sex, presented as box-and-whisker plots with individual data points. Overall (n = 28; gray), male (n = 19; blue), and female (n = 9; pink) distributions are shown for all 15 elements. Metals are arranged by concentration: high-concentration elements (Zn, Fe, Cu, Mn, Al; top row), mid-concentration elements (Cr, Se, Ni, Pb, Hg; middle row), and low-concentration elements (Co, As, Cd, V, Be; bottom row). Unadjusted Mann–Whitney *P* values for male vs. female comparisons are shown on each panel. No metals reached significance after Benjamini–Hochberg FDR correction. Chromium (p = 0.077) and iron (p = 0.085) showed borderline trends toward higher concentrations in males. The unequal group sizes (19 males vs. 9 females) limit statistical power.

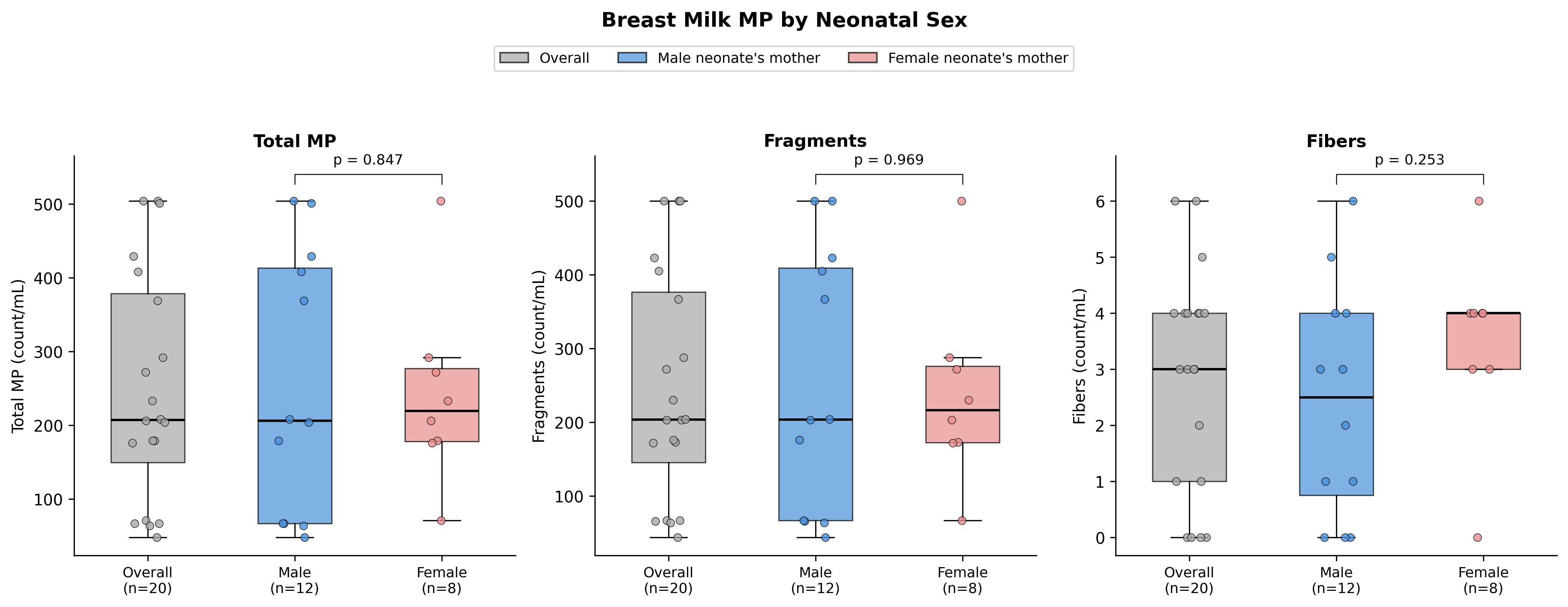

**Fig S6.** Breast milk microplastic counts (particles/mL) by neonatal sex, presented as box-and-whisker plots with individual data points. Overall (n = 20; gray), mothers of male neonates (n = 12; blue), and mothers of female neonates (n = 8; pink) distributions are shown for total MP, fragments, and fibers. No significant differences were observed between groups for any parameter (total MP: p = 0.847; fragments: p = 0.969; fibers: p = 0.253). The breast-milk pattern did not parallel the borderline sex difference observed in meconium (p = 0.045).

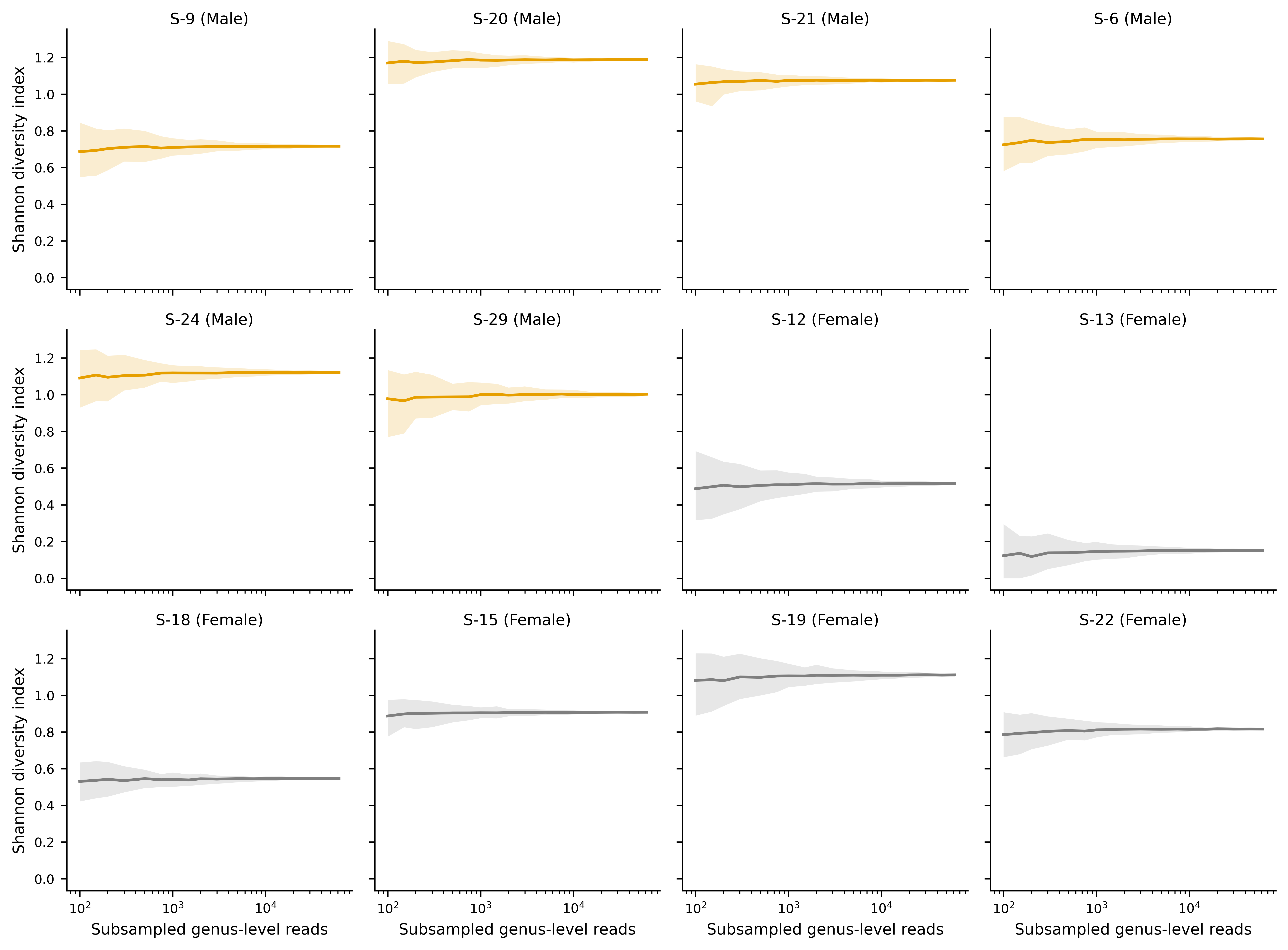

**Fig. S7. Subsampling-based sequencing-depth analysis supports robustness of genus-level Shannon diversity estimates.** Genus-level relative abundance profiles from the 12 neonatal meconium samples were repeatedly subsampled across increasing simulated read depths ranging from 100 to 61,899 genus-level reads. At each depth, 100 multinomial resampling iterations were generated from the Emu-estimated genus distribution for each sample and Shannon diversity was recalculated. Lines represent the mean Shannon diversity across iterations and shaded regions indicate the 95% simulation interval. Curves stabilized rapidly and remained essentially unchanged at higher simulated depths across all samples, indicating that the observed genus-level diversity estimates were unlikely to be driven primarily by insufficient sequencing depth. Because Emu produces estimated taxonomic abundance profiles rather than conventional ASV/OTU count tables, this analysis was used as a sequencing-depth robustness assessment rather than strict count-based rarefaction. Male and female samples are distinguished visually only and no group-level inference was performed in this analysis.

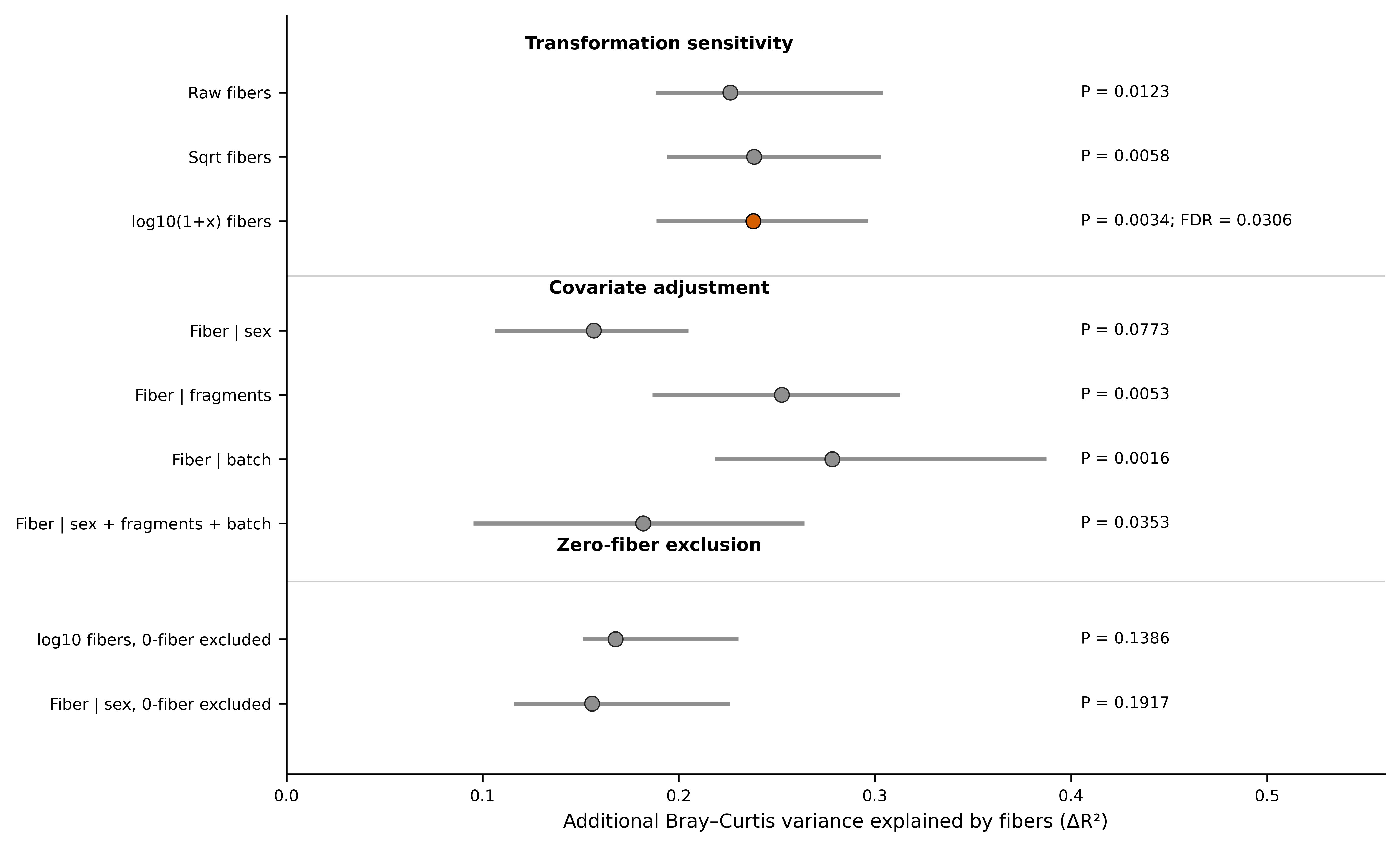

**Fig. S8. Sensitivity analyses of the exploratory association between microplastic fiber abundance and genus-level meconium community composition.** The association between fiber abundance and genus-level Bray-Curtis composition was evaluated across alternative fiber transformations, covariate-adjusted models, and analyses excluding the two samples in which no fibers were detected. Points represent the full-data additional variance attributable to fibers (ΔR²), and horizontal lines show the minimum-to-maximum ΔR² observed across leave-one-out analyses. The highlighted log10(1 + fiber abundance) model represents the primary fiber result from the exploratory metadata screen (R² = 0.238, P = 0.0034, FDR = 0.0306). Comparable effect sizes were observed using raw and square-root-transformed fiber abundance, and the association persisted after adjustment for fragment abundance and sequencing run/batch. In contrast, statistical support weakened after adjustment for neonatal sex (ΔR² = 0.157, P = 0.077) and after exclusion of the two zero-fiber samples (R² = 0.168, P = 0.139). The overall pattern therefore indicates a potentially meaningful but non-independent fiber-associated compositional gradient whose interpretation is limited by the small sample size, covariance with neonatal sex, and leverage of the zero-fiber observations.

**Supplementary tables**

**Table S1** Concentrations of reference material (SRM 1648a: Urban Dust for Trace Metals and other Constituents), and recoveries for metals

| **Metal** | **Unit** | **Certified Mass Fraction Value** | **Recovery Range (%)** |
| --- | --- | --- | --- |
| As | (mg/Kg) | 115 ± 3.9 | 93.09 - 98.45 |
| Al | % | 3.43 ± 0.13 | 94.53-104.34 |
| Cd | (mg/Kg) | 73.7 ± 2.3 | 96.11-105.32 |
| Cr | (mg/Kg) | 402 ± 13 | 94.49-99.12 |
| Cu | (mg/Kg) | 610 ± 70 | 95.43-103.29 |
| Ca | % | 5.84 ± 0.19 | 93.76-102.34 |
| Fe | % | 3.92 ± 0.21 | 91.82-98.76 |
| Pb | % | 0.655 ± 0.033 | 96.23-101.65 |
| Mg | % | 0.813 ± 0.012 | 97.43-108.32 |
| Mn | (mg/Kg) | 790 ± 44 | 98.50-103.23 |
| Ni | (mg/Kg) | 81.1 ± 6.8 | 96.45-102.02 |
| K | % | 1.056 ± 0.049 | 97.65-104.32 |
| Na | (mg/Kg) | 4240 ± 60 | 96.88-105.12 |
| Ti | (mg/Kg) | 4021 ± 86 | 94.87-99.11 |
| V | (mg/Kg) | 127 ± 11 | 95.73-100.88 |
| Zn | (mg/Kg) | 4800 ± 270 | 94.02-101.76 |

**Table S2:** Limits of Detection (LOD) and quantification (LOQ) for elements analysis in ICPMS

| **Elements** | **Unit** | **LOD** | **LOQ** |
| --- | --- | --- | --- |
| As | (µg/L) | 0.0124 | 0.124 |
| Se | (µg/L) | 0.011 | 0.111 |
| Pb | (µg/L) | 0.029 | 0.29 |
| Be | (µg/L) | 0.0012 | 0.013 |
| Cd | (µg/L) | 0.0052 | 0.053 |
| Co | (µg/L) | 0.012 | 0.13 |
| Cr | (µg/L) | 0.063 | 0.62 |
| Cu | (µg/L) | 0.051 | 0.49 |
| Mn | (µg/L) | 0.072 | 0.73 |
| Ni | (µg/L) | 0.063 | 0.61 |
| Zn | (µg/L) | 0.11 | 0.98 |
| V | (µg/L) | 0.0049 | 0.052 |
| Fe | (µg/L) | 0.12 | 1.02 |
| Al | (µg/L) | 0.078 | 0.81 |
| Hg | (µg/L) | 0.0013 | 0.012 |

**Precision:**

**Table S3:** Precision (CV%) for certified reference material and spiked-sample measurements.

| Elements | Precision (CV %) | | | |
| --- | --- | --- | --- | --- |
|  | CRM | | Spiked samples | |
|  | Intra-day (n=10) | Inter-day (n=30) | Intra-day (n=10) | Inter-day (n=30) |
| As | 2.5 | 3.4 | 3.9 | 4.8 |
| Se | 2.7 | 3.6 | 3.2 | 5.9 |
| Pb | 2.3 | 4.1 | 3.7 | 5.1 |
| Cd | 1.1 | 1.9 | 2.6 | 3.7 |
| Cr | 3.9 | 5.9 | 5.1 | 7.1 |
| Ni | 3.8 | 5.3 | 4.9 | 6.9 |
| Cu | 2.9 | 4.8 | 4.4 | 5.8 |
| Zn | 4.9 | 6.1 | 5.2 | 7.1 |
| Hg | 3.1 | 4.1 | 4.3 | 6.1 |
| Mn | 2.3 | 4.9 | 4.2 | 5.7 |
| Co | 2.2 | 3.1 | 3.4 | 4.9 |
| V | 1.9 | 2.7 | 3.1 | 3.8 |
| Be | BDL | BDL | 1.2 | 1.7 |

BDL = Below Detection Level

**Table S4:** Concentrations of reference material (SRM 955d: Toxic Metals and Metabolites in Frozen Human Blood), and recoveries for metals followed by microwave digestion process.

| **Metal** | **Unit** | **Certified Mass Concentration Values** | **Recovery Range (%)** |
| --- | --- | --- | --- |
| Lead (Pb) | (µg/dL) | Level 1- 1.480 ± 0.026  Level 2- 4.947 ± 0.085  Level 3- 42.13 ± 0.63 | 96.09 - 101.95 |
| Arsenic (As) | (µg/L) | Level 1- 5.31 ± 0.76  Level 2- 277.5 ± 4.8  Level 3- 774 ± 13 | 97.93-103.41 |
| Cadmium (Cd) | (µg/L) | Level 1- 0.326 ± 0.01  Level 2- 5.343 ± 0.082  Level 3- 10.50 ± 0.11 | 98.02-102.17 |
| Chromium (Cr) | (µg/L) | Level 1- 0.886 ± 0.069  Level 2- 2.012 ± 0.037  Level 3- 42.5 ± 1.3 | 95.87-100.87 |
| Cobalt (Co) | (µg/L) | Level 1- 0.384 ± 0.051  Level 2- 1.610 ± 0.057  Level 3- 31.91 ± 0.50 | 96.87-101.12 |
| Manganese (Mn) | (µg/L) | Level 1- 48.8 ± 1.3  Level 2- 51.2 ± 1.3  Level 3- 75.9 ± 2.0 | 95.96-101.45 |
| Mercury (Hg) Total | (µg/L) | Level 1- 1.373 ± 0.081  Level 2- 6.83 ± 0.33  Level 3- 55.3 ± 1.6 | 96.82-103.76 |
| Selenium (Se) | (µg/L) | Level 1- 206.6 ± 9.5  Level 2- 263 ± 16  Level 3- 749 ± 62 | 96.23-101.65 |

**Supplementary Table Captions (Supplementary materials 2)**

**Table S5.** Per-sample demographic characteristics, microplastic counts, and polymer identification for 30 neonates enrolled at Ad-Din Medical College Hospital, Dhaka, Bangladesh. Meconium MP counts are presented as raw values (per 0.5 g) and normalized values (per 1 g wet weight). Samples S8 and S27 were lost during processing. Samples exceeding 500 fragments per 0.5 g are recorded as "500+."

**Table S6.** Concentrations of 15 trace elements in neonatal meconium (n = 28) measured by ICP-MS, reported in µg/g dry weight. Samples S18 and S19 were excluded due to insufficient volume.

**Table S7.** Microplastic contamination in two commercially available infant formula products, including fragment and fiber counts per mL, polymer identification, and particle size measurements. Data are exploratory (n = 2).

**Table S8.** Paired mother–neonate microplastic comparison for 19 valid pairs. Meconium MP counts are reported as particles per gram wet weight; breast milk counts are as particles per mL. Polymer types identified by Raman spectroscopy are listed for each matrix.

**Table S9.** Pairwise Spearman's rank correlation matrix for all 15 trace elements in neonatal meconium (n = 28). Pairs are sorted by ascending p-value. Correlations with unadjusted p < 0.05 are shown in bold as nominal descriptive associations. No multiple-testing correction was applied.

**Table S10.** Complete results of demographic association analyses. (A) Neonatal sex vs. meconium MP counts (Mann–Whitney U, n = 28). (B) Gestational age vs. meconium MP counts (Spearman, n = 28). (C) Term (≥37 weeks; n = 17) vs. late preterm (<37 weeks; n = 11) meconium MP counts (Mann–Whitney U). (D) Neonatal sex vs. heavy metal concentrations (Mann–Whitney U, n = 28; 19 males, 9 females) with BH-FDR correction. (E) Gestational age vs. heavy metal concentrations (Spearman, n = 28) with BH-FDR correction. Se and Cu were the only metals reaching FDR-corrected significance.

**Table S11.** Exploratory associations between available clinical and exposure variables and genus-level neonatal meconium microbiome diversity and composition. Shannon diversity was evaluated using exact permutation comparisons for binary variables and permutation-based Spearman correlations for continuous variables, while overall community composition was evaluated by PERMANOVA using genus-level Bray-Curtis dissimilarities. Effect estimates, Benjamini-Hochberg false-discovery-rate-adjusted P values, and leave-one-out sensitivity ranges are shown for the 12 microbiome samples. Neonatal sex showed a moderate but non-significant association with both Shannon diversity and community composition, whereas total microplastic abundance, fragment abundance, and categorical microplastic burden showed little evidence of association with Bray-Curtis structure. Fiber abundance was the only screened variable associated with community composition after FDR correction, explaining approximately 24% of Bray-Curtis variation. Dispersion P values are shown for binary comparisons as a diagnostic of within-group heterogeneity. These findings are exploratory and should be interpreted in light of the small sample size and limited independence among some covariates.

| **Predictor** | **Contrast / transformation** | **Shannon effect** | **Shannon P** | **Shannon FDR** | **LOO Shannon effect, median [min-max]** | **PERMANOVA R² (pseudo-F)** | **PERMANOVA P** | **PERMANOVA FDR** | **LOO R², median [min-max]** | **Dispersion P** |
| --- | --- | --- | --- | --- | --- | --- | --- | --- | --- | --- |
| Neonatal sex | Male vs Female | Δmean = 0.302; Cliff's δ = 0.556 | 0.0844 | 0.2386 | 0.289 [0.197-0.389] | 0.165 (1.974) | 0.0844 | 0.3799 | 0.173 [0.137-0.231] | 0.1255 |
| Term status | Preterm vs Term | Δmean = -0.003; Cliff's δ = 0.000 | 0.9913 | 0.9949 | 0.005 [-0.139-0.058] | 0.051 (0.536) | 0.8030 | 0.9034 | 0.055 [0.037-0.097] | 0.5108 |
| Feeding | Mixed vs exclusive breastfeeding | Δmean = -0.217; Cliff's δ = -0.630 | 0.3182 | 0.5727 | -0.208 [-0.308--0.144] | 0.035 (0.360) | 0.9227 | 0.9227 | 0.039 [0.027-0.092] | 0.6591 |
| Baby product | Katha vs Diaper | Δmean = 0.085; Cliff's δ = 0.086 | 0.6591 | 0.8754 | 0.073 [-0.102-0.171] | 0.078 (0.841) | 0.5240 | 0.8293 | 0.083 [0.061-0.123] | 0.9457 |
| MP category | Low vs High | Δmean = -0.287; Cliff's δ = -0.500 | 0.1061 | 0.2386 | -0.262 [-0.375-0.301] | 0.067 (0.716) | 0.6450 | 0.8293 | 0.074 [0.055-0.105] | 0.2597 |
| Gestational age | Continuous, weeks | Spearman ρ = 0.134 | 0.6809 | 0.8754 | 0.105 [-0.055-0.323] | 0.089 (0.977) | 0.4379 | 0.8293 | 0.092 [0.056-0.171] |  |
| Total MP abundance | Continuous, log10(particles/g) | Spearman ρ = 0.543 | 0.0771 | 0.2386 | 0.563 [0.405-0.651] | 0.068 (0.725) | 0.6250 | 0.8293 | 0.075 [0.051-0.097] |  |
| Fragment abundance | Continuous, log10(fragments/g) | Spearman ρ = 0.575 | 0.0562 | 0.2386 | 0.584 [0.447-0.671] | 0.070 (0.750) | 0.5992 | 0.8293 | 0.077 [0.059-0.100] |  |
| Fiber abundance | Continuous, log10(1 + fibers/g) | Spearman ρ = -0.004 | 0.9949 | 0.9949 | -0.014 [-0.150-0.220] | 0.238 (3.123) | 0.0034 | 0.0306 | 0.245 [0.189-0.297] |  |

***Notes:*** *For binary predictors, Shannon effect estimates are differences in group means (first group minus second group), with Cliff's delta included as an additional rank-based effect size. For continuous predictors, Shannon associations are Spearman rank correlations. Community composition was evaluated using genus-level Bray-Curtis dissimilarities. Benjamini-Hochberg FDR correction was applied separately across the exploratory Shannon and PERMANOVA metadata screens. Leave-one-out (LOO) values summarize analyses repeated after sequential omission of each sample. Total MP and fragment abundance were log10-transformed; fiber abundance was analyzed as log10(1 + fibers/g) to retain zero-fiber observations. Dispersion P values are reported only for binary predictors. S-20 and S-21 represented lower-bound particle counts at the analytical counting ceiling.*

**Table S12.** Sensitivity analyses of the exploratory association between microplastic fiber abundance and genus-level neonatal meconium community composition. The primary fiber association was evaluated across alternative transformations and after adjustment for neonatal sex, fragment abundance, and sequencing run, with additional analyses excluding the two samples in which no fibers were detected. ΔR² represents the additional fraction of total Bray-Curtis variation attributable to fibers relative to the corresponding reduced model, while partial R² represents the fraction of residual variation attributable to fibers after adjustment for included covariates. Leave-one-out ranges summarize the stability of effect estimates after sequential omission of individual samples. The fiber-associated compositional signal was similar across transformations and persisted after adjustment for fragment abundance and sequencing run, but was attenuated after adjustment for neonatal sex and after exclusion of the two zero-fiber samples. The result therefore supports fiber abundance as a hypothesis-generating correlate of community composition rather than evidence of an independent or causal microbiome effect.

| **Analysis** | **Model** | **n** | **ΔR² total** | **Partial R²** | **P** | **FDR** | **LOO ΔR², median [min-max]** | **LOO partial R², median [min-max]** |
| --- | --- | --- | --- | --- | --- | --- | --- | --- |
| Transformation sensitivity | Raw fiber abundance | 12 | 0.226 | 0.226 | 0.0123 |  | 0.239 [0.189-0.304] | 0.239 [0.189-0.304] |
| Transformation sensitivity | Square-root fiber abundance | 12 | 0.238 | 0.238 | 0.0058 |  | 0.249 [0.194-0.303] | 0.249 [0.194-0.303] |
| Transformation sensitivity | log10(1 + fiber abundance) | 12 | 0.238 | 0.238 | 0.0034 | 0.0306 | 0.245 [0.189-0.297] | 0.245 [0.189-0.297] |
| Covariate adjustment | Fiber adjusted for neonatal sex | 12 | 0.157 | 0.188 | 0.0773 |  | 0.166 [0.106-0.205] | 0.199 [0.138-0.252] |
| Covariate adjustment | Fiber adjusted for fragment abundance | 12 | 0.253 | 0.271 | 0.0053 |  | 0.262 [0.186-0.313] | 0.282 [0.207-0.341] |
| Covariate adjustment | Fiber adjusted for sequencing run | 12 | 0.278 | 0.314 | 0.0016 |  | 0.282 [0.218-0.387] | 0.325 [0.252-0.442] |
| Covariate adjustment | Fiber adjusted for sex + fragments + sequencing run | 12 | 0.182 | 0.282 | 0.0353 |  | 0.184 [0.095-0.264] | 0.291 [0.177-0.453] |
| Zero-fiber exclusion | log10(1 + fiber abundance), zero-fiber samples excluded | 10 | 0.168 | 0.168 | 0.1386 |  | 0.175 [0.151-0.230] | 0.175 [0.151-0.230] |
| Zero-fiber exclusion | Fiber adjusted for sex, zero-fiber samples excluded | 10 | 0.156 | 0.177 | 0.1917 |  | 0.161 [0.116-0.226] | 0.181 [0.139-0.261] |

***Notes:*** *For unadjusted models, ΔR² and partial R² are identical. For the primary unadjusted log10(1 + fiber abundance) model, the reported P value and FDR are from the exploratory metadata screen; the remaining P values are from targeted sensitivity analyses. Adjusted-model P values were generated using residual permutation of the tested fiber predictor relative to the included covariates. FDR correction was not reapplied across the targeted sensitivity models. Sequencing run was included only as a technical sensitivity variable and was not treated as a biological predictor. The fully adjusted model contains several predictors for only 12 samples and should therefore be interpreted cautiously. Zero-fiber exclusion analyses used n = 10.*
